# Measuring patient centeredness with German language Patient-Reported Experience Measures (PREM) – a systematic review and qualitative analysis according to COSMIN

**DOI:** 10.1101/2022.02.04.22270374

**Authors:** André L. Mihaljevic, Colette Dörr-Harim, Eva Kalkum, Guido Strunk

**Affiliations:** Department of General and Visceral Medicine, University Hospital Ulm, Albert-Einstein-Allee 23, 89081 Ulm, Germany; Clinical Trial Centre, Department of Surgery (ulmCARES), University Hospital Ulm, Albert-Einstein-Allee 23, 89081 Ulm, Deutschland; Study Centre of the German Society of Surgery, Im Neuenheimer Feld 130.3, 69120 Heidelberg, Germany; Complexity-Research, Schönbrunner Str. 32/3/20, A-1050 Wien, Austria

**Keywords:** patient-reported experience measure, patient-reported outcome measure, patient-centredness, patient outcome assessment

## Abstract

**Background:** Patient centeredness is an integral part of the quality of care. Patient-reported experience measures (PREMs) are assumed to be an appropriate tool to assess patient-centredness. An evaluation of German-speaking PREMs is lacking.

**Objective:** To perform a systematic review and qualitative analysis of psychometric measurement qualities of German-speaking PREMs.

**Methods:** A systematic literature search was performed in Medline, PsycInfo, CINHAL, Embase, Cochrane database (last search 9^th^ November 2021) for studies describing generic, surgery-or cancer-specific PREMs. Subsequently, questionnaires that were not in German or for which no German translation exists were excluded. Furthermore, questionnaires, that are for healthcare professionals rather than patients as well as disease-specific questionnaires were excluded. Baseline data for all studies was extracted by two independent reviewers. Psychometric measurement qualities of the questionnaires were assessed according to the COSMIN guidelines.

**Results:** In total, 3457 abstracts were screened, of which 3345 were excluded. The remaining 112 articles contained 51 PREMs, of which 12 were either developed in or translated into German: 8 generic (NORPEQ, PPE-15, PEACS, HCAHPS, QPPS, DUQUE, PEQ-G, Schoenfelder et al.), 4 cancer-specific (EORTC IN-PATSAT32, PSCC-G, Danish National Cancer Questionnaire, SCCC) and no surgery-specific PREM. None of the PREMs covered all 16 domains patient-centeredness. Overall rating of structural validity was adequate only for PEACS and HCAHPS. High ratings for internal consistency were given for NORPEQ, Schoenfelder et al., PSCC-G and the SCCC. Cross-cultural validity for translated questionnaires was adequate only for the PSCC-G, while reliability was adequately assessed only for the EORTC IN-PATSAT32. Due to a lack of measurement gold standard and minimal important change, criterion validity and measurement invariance could not be assessed Most PREMs showed the expected outcomes for hypotheses testing for construct validity.

**Conclusion:** None of the PREMs has been fully evaluated in German. Based on current evidence, the EORTC IN-PATSAT32 and PSCC-G can be recommended for cancer patients, while HCAHPS, NORPEQ, PPE-15 and PEACS might be used as generic PREMs. Compared to other languages PREM development in German is rudimentary and no comprehensive PREM system is currently available.

**Registration:** PROSPERO CRD42021276827

**Funding:** None

## Introduction

Improving the patient centredness (PC) of healthcare has been a main objective of healthcare politics over the last decades, including German-speaking countries [1, 2]. PC has been defined as one of six domains of the quality of care by the Institute of Medicine (IOM), next to safety, effectiveness, timeliness, efficiency and equitability [3] (S1 Fig). However, patients frequently experience a lack of PC in many fields of healthcare [4].

Furthermore, the dimensions of PC have not been clearly defined and several models have been proposed in the past (S2 Table). A systematic review has identified 15 dimensions of PC [5]: patient as a unique person, biopsychosocial perspective, essential characteristics of the clinician, patient involvement in care, involvement of family and friends, physical support, emotional support, clinician-patient communication, patient empowerment, patient Information, access to care, integration of medical and non-medical care, coordination and continuity of care, teamwork and teambuilding, clinician-patient relationship. The influential Picker model contains an additional dimension termed “effective treatment by trustworthy and qualified personnel” [6] (S2 Table).

Several methods have been proposed to measure PC in clinical practice [7]. Assessment via questionnaires termed *Patient-Reported Experience Measures* (PREM) is most frequent as they permit a standardized appraisal of PC. PREMs aim to measure PC via the experience of patients in a certain healthcare context. Depending on this healthcare context four different categories of PREMs can be distinguished, although they partially overlap:

a. *generic PREMs* measure general aspects of PC and can be applied across multiple healthcare settings and disciplines.
b. *discipline-specific PREMs* assess the PC within a certain discipline, e.g. surgery or internal medicine.
c. *healthcare pathway-specific PREMs* measure the PC across a specific healthcare pathway e.g., cancer care and thus across disciplines and healthcare settings.
d. *disease-specific PREMs* aim to measure the PC of healthcare for patients with a specific disease, e.g., breast cancer.

According to Bull et al. [8] PREMs (a) enable patients to reflect on their care experience; (b) provide patient-level information to drive service quality improvement; (c) serve as quality indicators for public reporting and benchmarking. PREMs need to be distinguished from *Patient-Reported Outcome Measures* (PROM). PROMs measure medical outcomes, rather than dimensions of PC and therefore, address different domains of healthcare in the IOM quality of care model (S2). Like PROMs, however, PREMs need to have adequate psychometric measurement properties (e.g., validity, reliability) to accurately assess PC. However, non-standardized PREMs or PROMs are frequent [9] and constitute a waste of resources [10]. The COSMIN (COnsensus-based Standards for the selection of health Measurement INstruments) group has defined internationally accepted guidelines for assessing psychometric properties of patient-reported measures [11].

Contrary to other languages no comprehensive review is available for German PREMs. Furthermore, the psychometric properties of German PREMs have not been evaluated yet. Therefore, the aim of the study is to perform a systematic review and qualitative analysis of generic, surgery-specific and cancer-care specific German PREMs using the internationally accepted COSMIN guidelines [11].

## Material and Methods

This systematic review is reported according to current PRISMA guidelines [12]. A PRISMA checklist is attached (S3 Table). The review has been registered (PROSPERO CRD42021276827). No funding has been received for this work. A review protocol had been written prior to data extraction.

### Eligibility criteria

Studies that described or analysed the development, validation, or measurement of generic, surgery-specific or cancer healthcare pathway-specific PREMs were included in this systematic review. Included studies needed to be published in a peer-reviewed journal. During the initial search no language limitations were set. The following exclusion criteria were set:

1. Studies that described general satisfaction questionnaires that did not cover aspects of patient experience.
2. Studies measuring patient expectation rather than patient experience
3. Studies that measured single aspects of patient-centeredness and were thus no multidimensional PREMs
4. Studies describing questionnaires for physicians or proxies and not for patients themselves (i.e., were not patient-reported)
5. Studies containing PREMs that were not generic, surgery-specific or cancer healthcare pathway-specific. Therefore, PREMs for a specific disease were excluded.

### Information sources

The following information sources were searched:

a. EBM Reviews -Cochrane Database of Systematic Reviews 2005 to November 9, 2021, EBM Reviews -Health Technology Assessment 4th Quarter 2016
b. Embase 1980 to 2021 Week 40
c. Ovid MEDLINE(R) ALL 1946 to November 9, 2021
d. APA PsycInfo 1806 to November Week 2 2021
e. CINHAL November 9, 2021

The last search was performed on 9^th^ November 2021. An existing literature review on PREMs by Bull et al. was used as template [8]. This publication searched PREMs until 31^st^ March 2018. Therefore, our search was limited to 2018 till November 2021. The results by Bull et al. were included in this systematic review.

### Search

The search algorithm is described in detail in supplement 4 (S4 Text). It is an adaptation of the search algorithm described by Bull et al. [8]. Additional studies were identified by reference searching and full text reading.

### Study selection

The references of all generic, surgery-specific or cancer healthcare pathway-specific PREMs were imported into the citation program Zotero (www.zotero.org; Version 5.0.96.2). Duplicates were identified and merged either with the find duplicates function in Zotero or by hand. Titles and abstracts of all articles were read by two reviewers (AMi, CDH) and those studies not fulfilling eligibility criteria were removed. In a next step, the fulltext articles of all remaining studies were read, to decide which articles fulfil eligibility criteria. Fulltext as well as references were screened to identify additional PREMs. For all non-German PREMs fulfilling the eligibility criteria additional searches were performed in the above-mentioned databases to identify German translations. In addition, Google and google scholar were search with the name of the PREM in combination with “German translation” or “cross-cultural validation” to identify German translations. Only PREMs developed in German or for which a German language translation existed were considered for further analyses.

### Data collection process

Data was extracted on prespecified forms by two reviewers (AMi, CDH). The following data items were collected: author, year, journal, PREM acronym, country of origin, description of the PREM, method and timepoint of PREM collection, number of questions, PREM domains according to original article, presence of a German version, free text commentary.

### Assessment of psychometric properties and data synthesis

Psychometric properties were used according to the COnsensus-based Standards for the selection of health Measurement INstruments (COSMIN) terminology [13]. Qualitative analysis of psychometric properties was done in according to current COSMIN guidelines [14, 15] (www.cosmin.nl). The evaluation of content validity was done according to Terwee et al. [11]. We used the 16 PC dimensions described in S1 to assess content validity. The following psychometric properties were analysed: (1) content validity; (2) structural validity; (3) internal consistency; (4) measurement invariance / cross-cultural validity; (5) reliability; (6) measurement error; (7) criterion validity; (8) hypothesis testing for construct validity. Table 1 shows the COMSIN assessment criteria used in this study. For overall evaluation “+” marks adequate, “-“ inadequate and “?” unclear psychometric properties. A detailed description of the methods can be found in S5 Text.

**Table 1.**
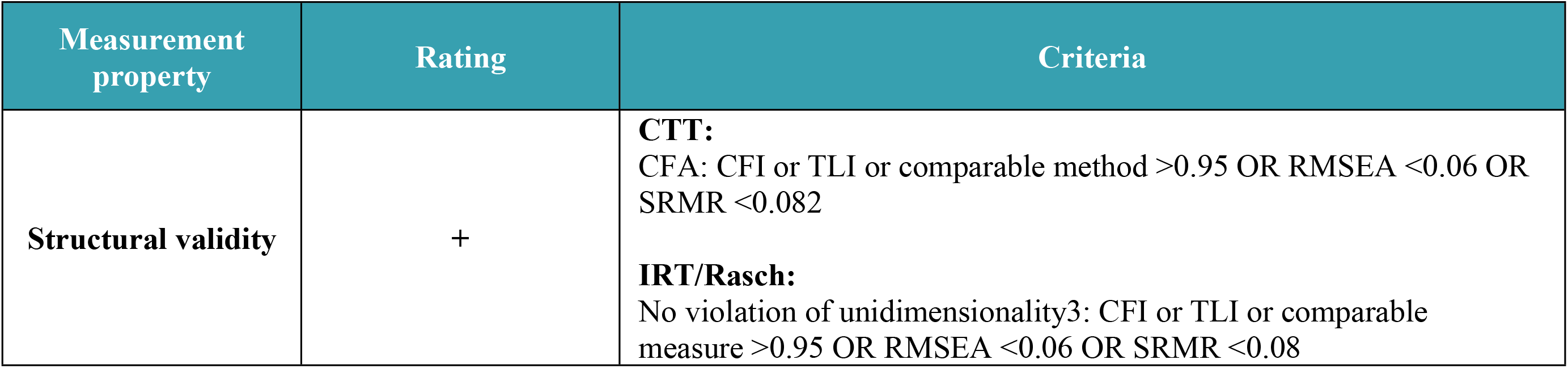
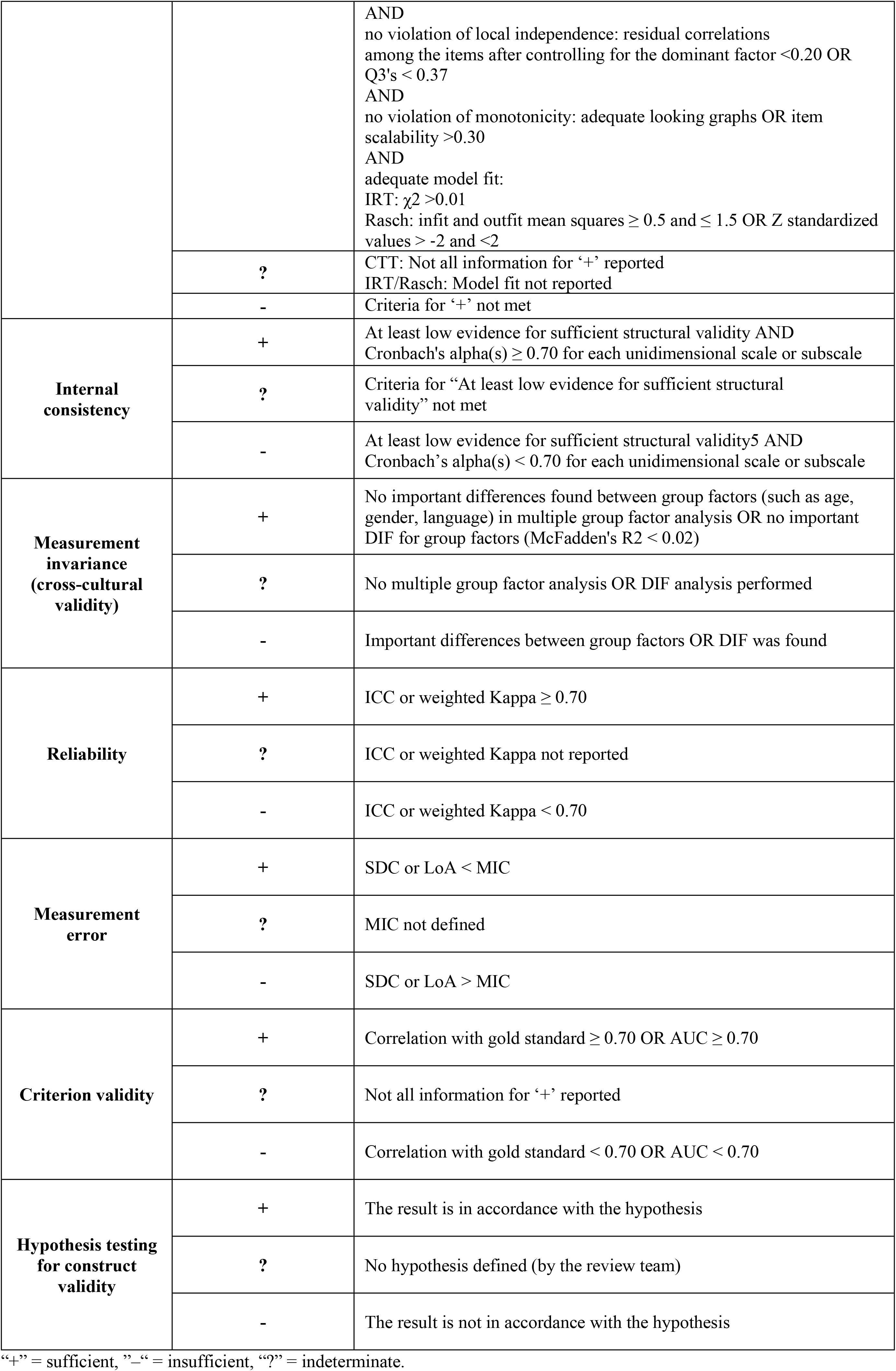
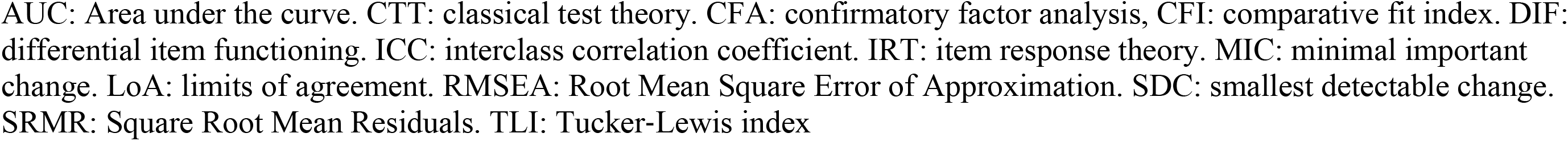
Criteria for overall assessment of psychometric properties according to COSMIN (Mokkink et al. 2018, COSMIN User Manuel).

## Results

### Study selection and study characteristics

By database searching 3,646 articles were identified. An additional 55 potential studies were found by hand-search and 22 PREMs were added through the systematic review by Bull et al. [8] (Fig 1). After screening abstracts and titles, 112 fulltext articles remained of which 51 articles fulfilled all eligibility criteria. Twelve of these 51 articles described German PREMs [16–27] (Table 2). Details about the 39 non-German PREMs can be found in S6 Table. Of the 12 German-language PREMs, eight were generic [16,18–20,24–27] and four were cancer-care specific PREMs [17,21–23]. No surgery-specific German-language PREM was identified. One of the German-language PREMs *(Quality from the Patients’ Perspective, QPP)* is available only in its short-form *(short-form, QPPS)* [28]. The PREMs are either translations into German [16–22] or were developed in German [23–26]. Study characteristics can be found in Table 2.

**Fig 1.**
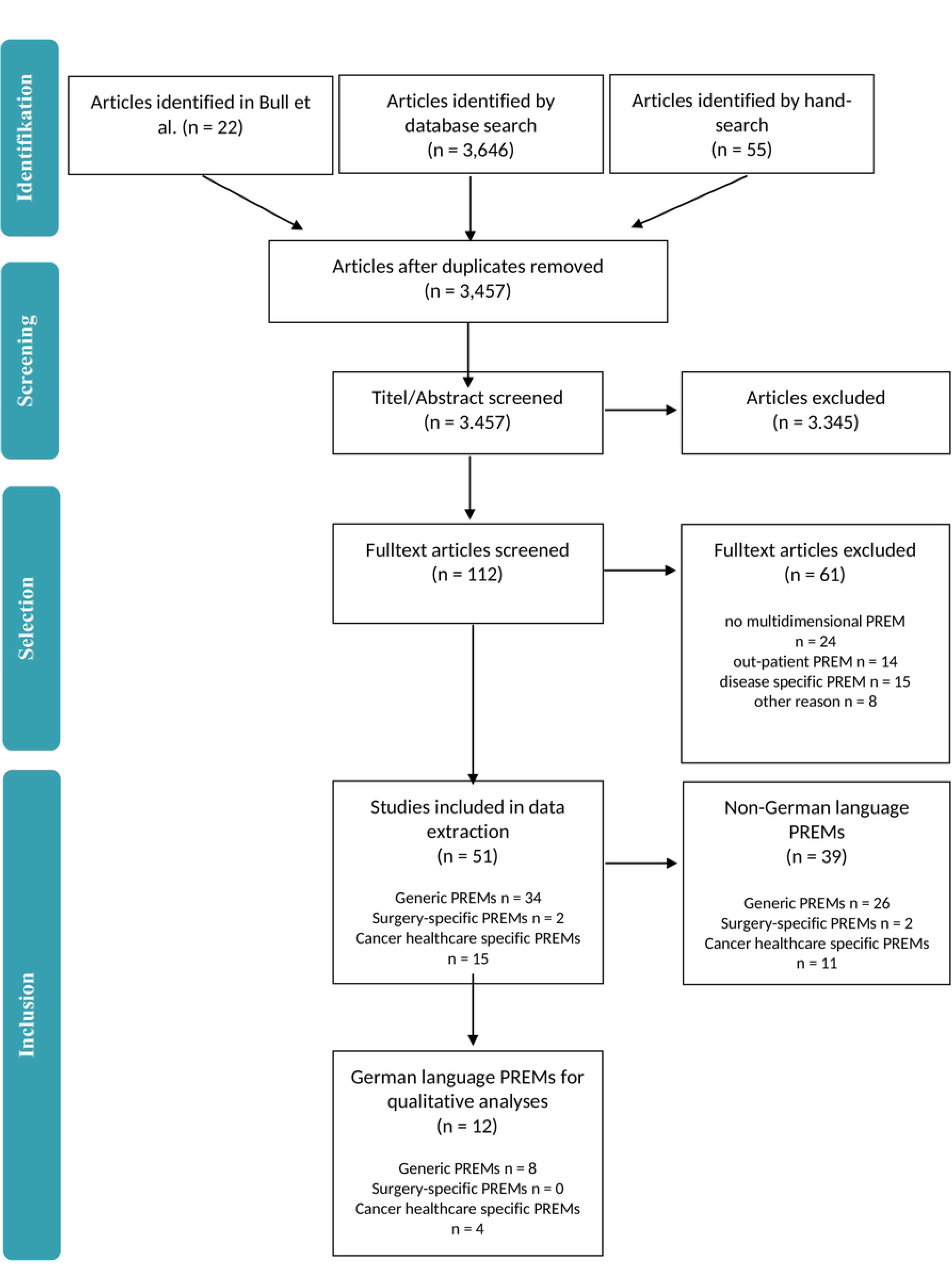
PRISMA Flow-chart of included studies.

**Table 2.**
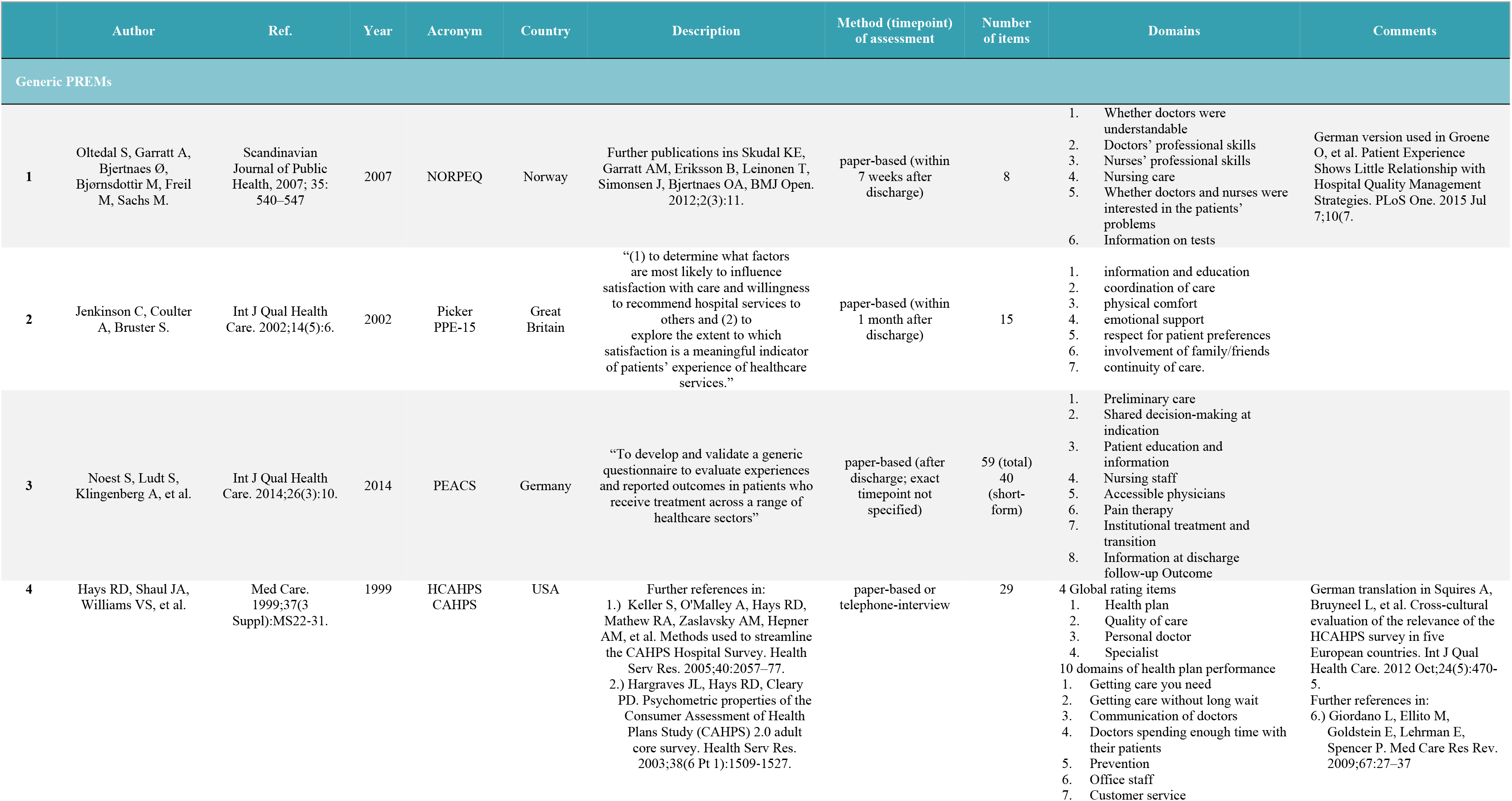
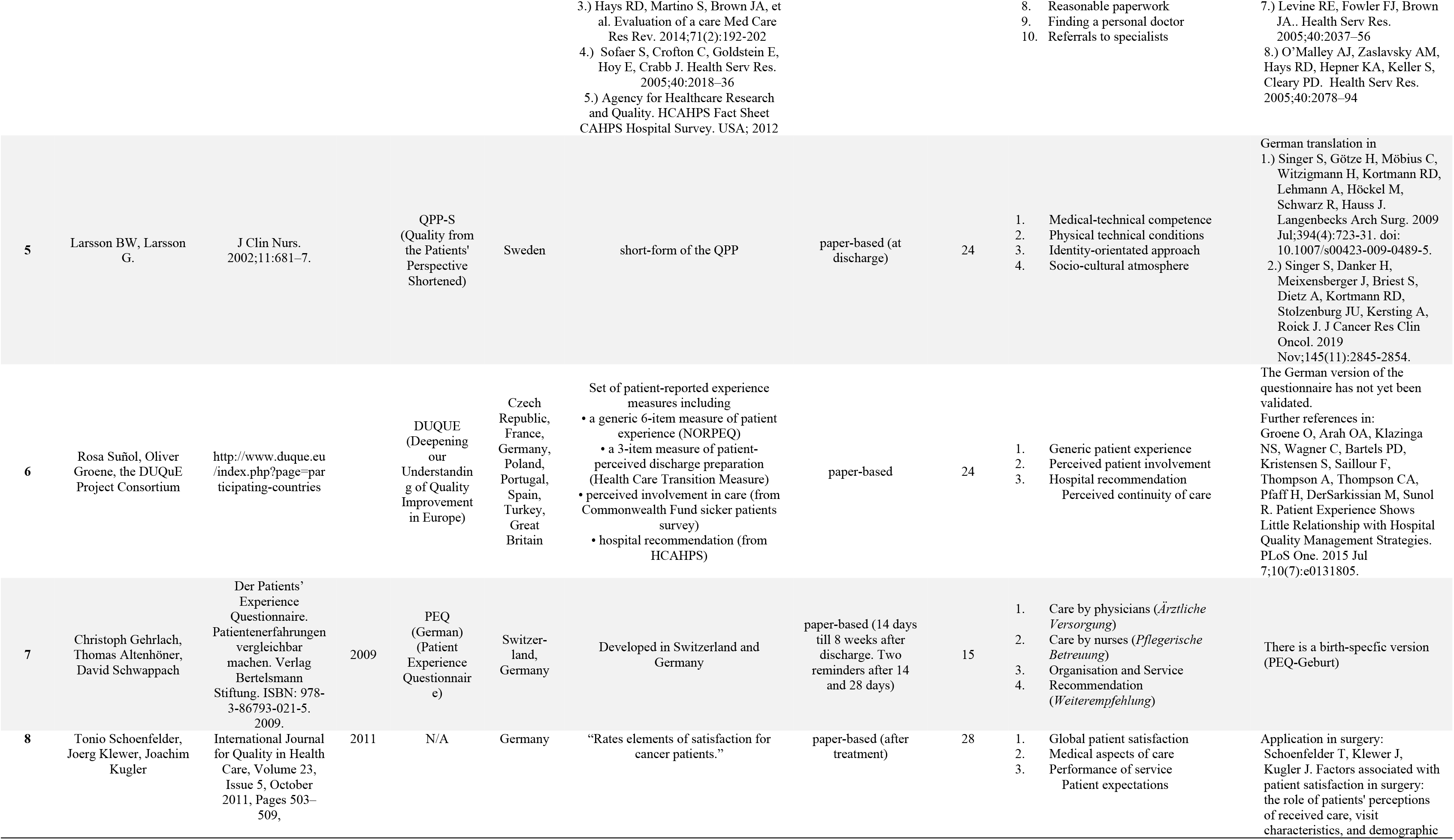
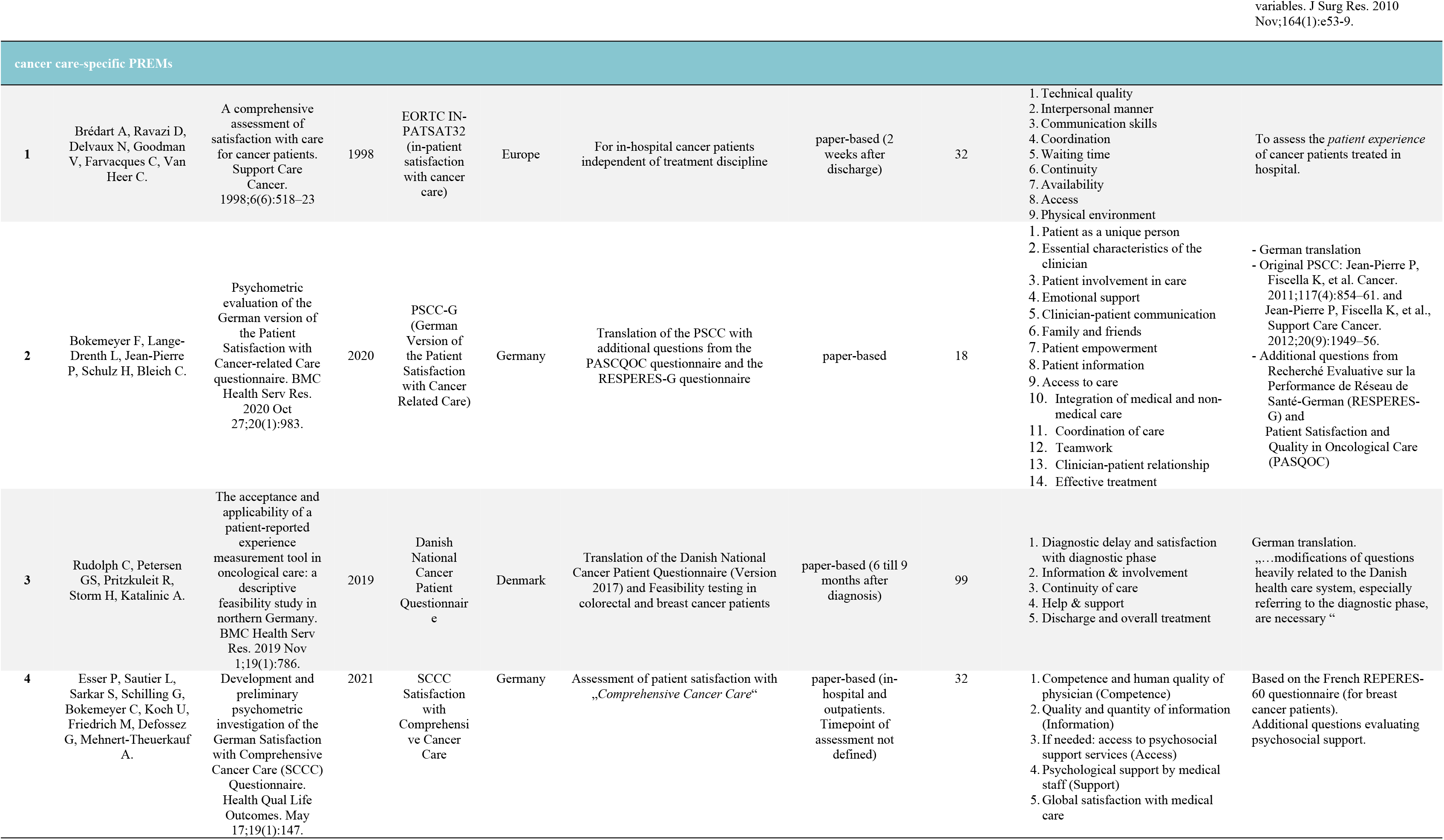
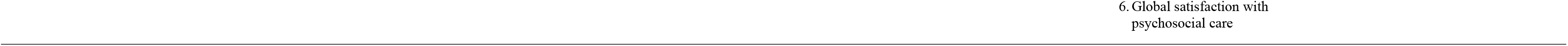
Study characteristics of German-language Patient-Reported Experience Measures (PREMs).

### Content validity

Content validity was evaluated using he 16 dimensions of PC outlined in the introduction (Table 3). None of the questionnaires covers all aspects of PC. An overview of the methodological quality of studies analyzing the content validity according to COSMIN can be found in Table 4. Alle 12 included PREMs exhibit sufficient relevance. Because of the lack of content dimensions all questionnaires have deficits in *comprehensiveness* (Table 4). Understandability of questionnaires is adequate in most cases. However, the methodological quality of content validity studies varies widely from low (Schoenfelder, DUQUE) to high (Picker, HCAHPS, PEQ, EORTC IN-PATSAT32, PSCC-G) (Table 4).

**Table 3.** Overview of content domains of German-language PREMs.

|  | PREM Domain | HCAHPS | PEACS | PPE-15 | NORPEQ | QPP* | QPPS | DUQUE | PEQ<br>(German) | Schoen-<br>felder et al. | EORTC<br>IN-<br>PATSAT32 | PSCC-G | Danish<br>National<br>CPQ | SCCC |
| --- | --- | --- | --- | --- | --- | --- | --- | --- | --- | --- | --- | --- | --- | --- |
| 1 | patient as a unique person | - | - | - | - | + | + | - | + | - | + / - | + | + | + |
| 2 | biopsychosocial perspective | - | - | - | - | + / - | - | - | - | + | - | - | - | + |
| 3 | essential characteristics of the clinician | - | - | - | + / - | - | - | + / - | - | + | + | + | + | + |
| 4 | patient involvement in care | - | + | + | + | + | + / - | + | + | + | - | + / -<sup>4</sup> | + | + |
| 5 | involvement of family and friends | - | + | + | - | + |  | + | - | - | - | + / -<sup>4</sup> | - | - |
| 6 | physical support | + | + | + | +/- | + | + | - | - | - | + |  | + | + |
| 7 | emotional support | - | + | + | +/- | + / - | + / - | + | - | - | + | + / - | + | + |
| 8 | clinician-patient communication | + | + | + | + | + / - | + / - | + | - | + | + | + | + | + |
| 9 | patient empowerment | - | + | - | - | + / - | - | + | - | - | - | + | - | + |
| 10 | patient Information | + | + | - | + | + | + / - | + | + | + | + | + / -<sup>4</sup> | + | + |
| 11 | access to care | - | + | - | - | - | - | - | + | - | + / - | + | - | + |
| 12 | integration of medical and non-medical care | - | + | - | - | - | - | - | - | - | - | + | - | + / - |
| 13 | coordination and continuity of care | +/- | + | + | - | - | - | + / - | + / - | + | - | + | + | + / - |
| 14 | teamwork and teambuilding | - | + | - | - | - | - | - | - | - | + / -<br>(only nursing care) | +<sup>1</sup> | - | - |
| 15 | clinician-patient relationship | +/- | + | - | +/- | + | + / - | + / - | + | - | + | + | + | + |
| 16 | effective treatment | - | + | - | + / - | + | + | - | + | + | + | + | - | + / - |

**Table 4.**
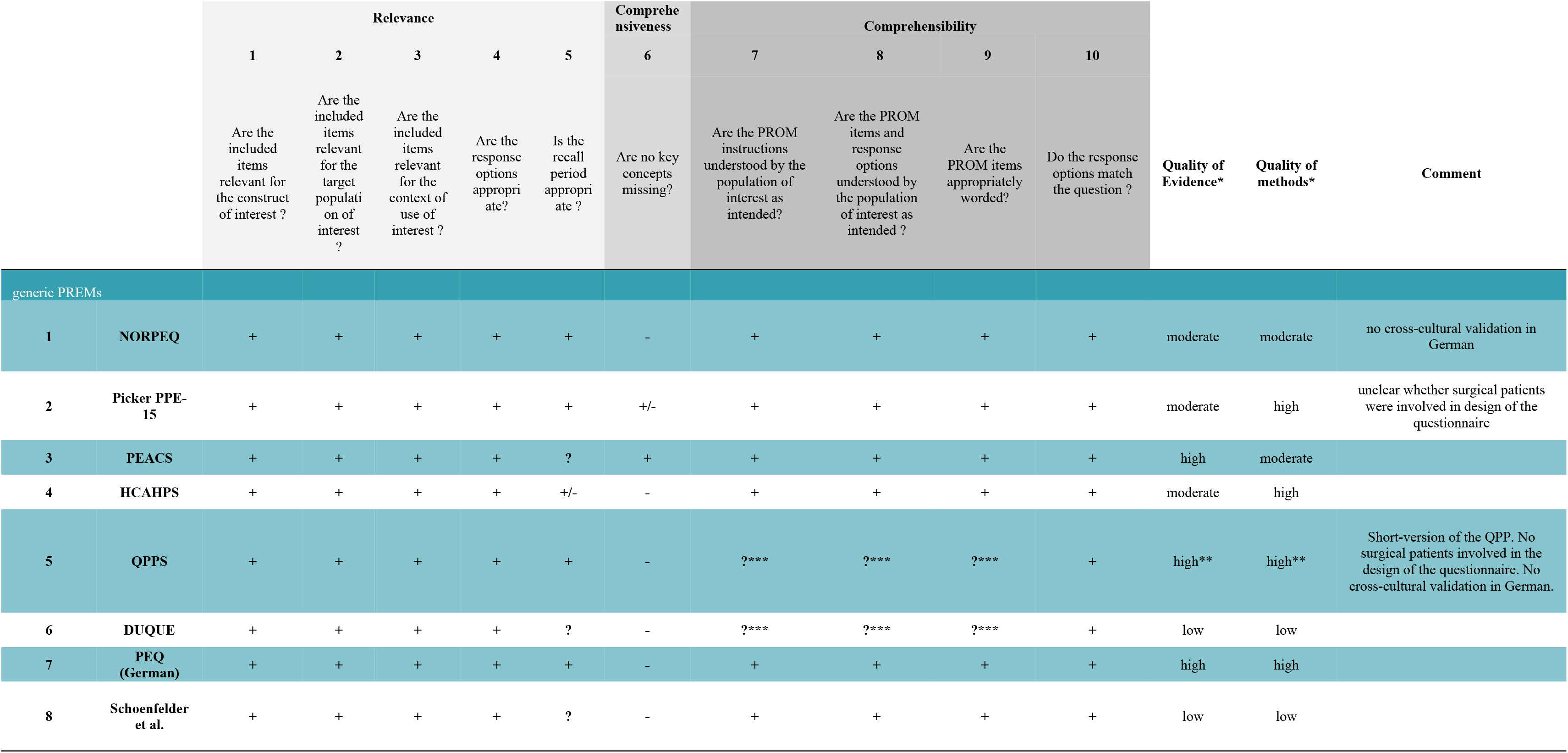
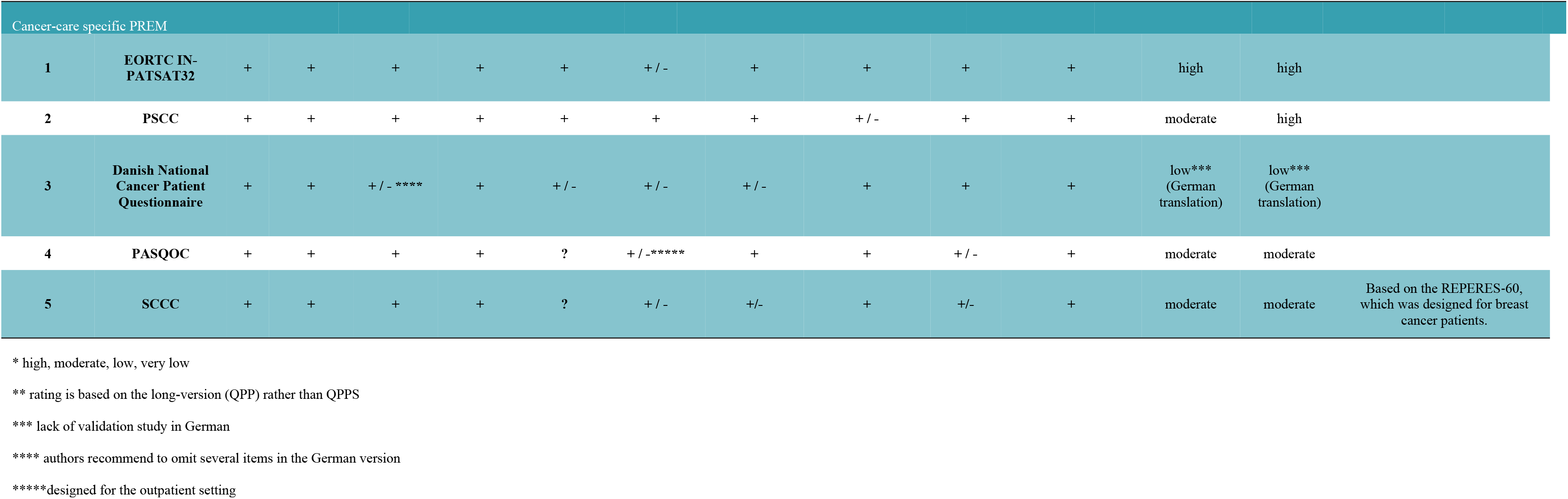
Rating of content validity of German PREMs according to COSMIN.

### Structural validity

“Structural validity refers to the degree to which the scores of a PROM (or PREM) are an adequate reflection of the dimensionality of the construct to be measured” [11]. A summary table of the findings on structural validity can be found in Table 5. Not all necessary data on confirmatory factor analyses according to current COSMIN guidelines were available to rate the NORPEQ or the questionnaire by Schoenfelder et al. (Table 5). The PPE-15 received an insufficient rating for structural validity due to the inadequate design of the underlying studies [16]. Similarly, the SCC showed insufficient structural validity. No data on structural validity could be obtained for the German PEQ neither in the original publication nor in additional studies [25]. The same was true for the Danish National Cancer Questionnaire. PEACS and HCAHPS received a sufficient rating (Table 5). For the EORTC INPAT-SAT32 there is a metanalysis summarizing the data on structural validity [29]. There is a detailed analysis of structural validity for the PSCC-G [22], which shows mixed results.

**Table 5.**
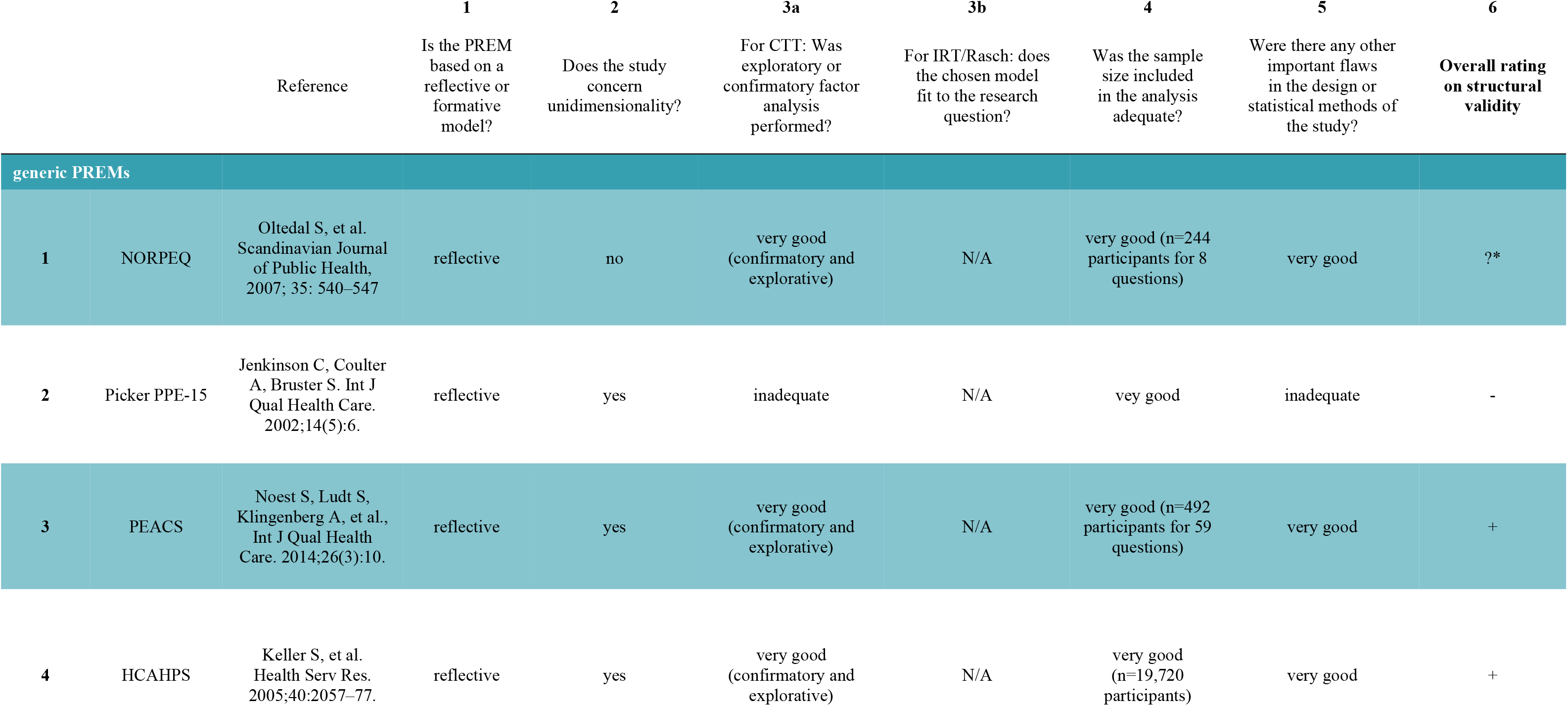
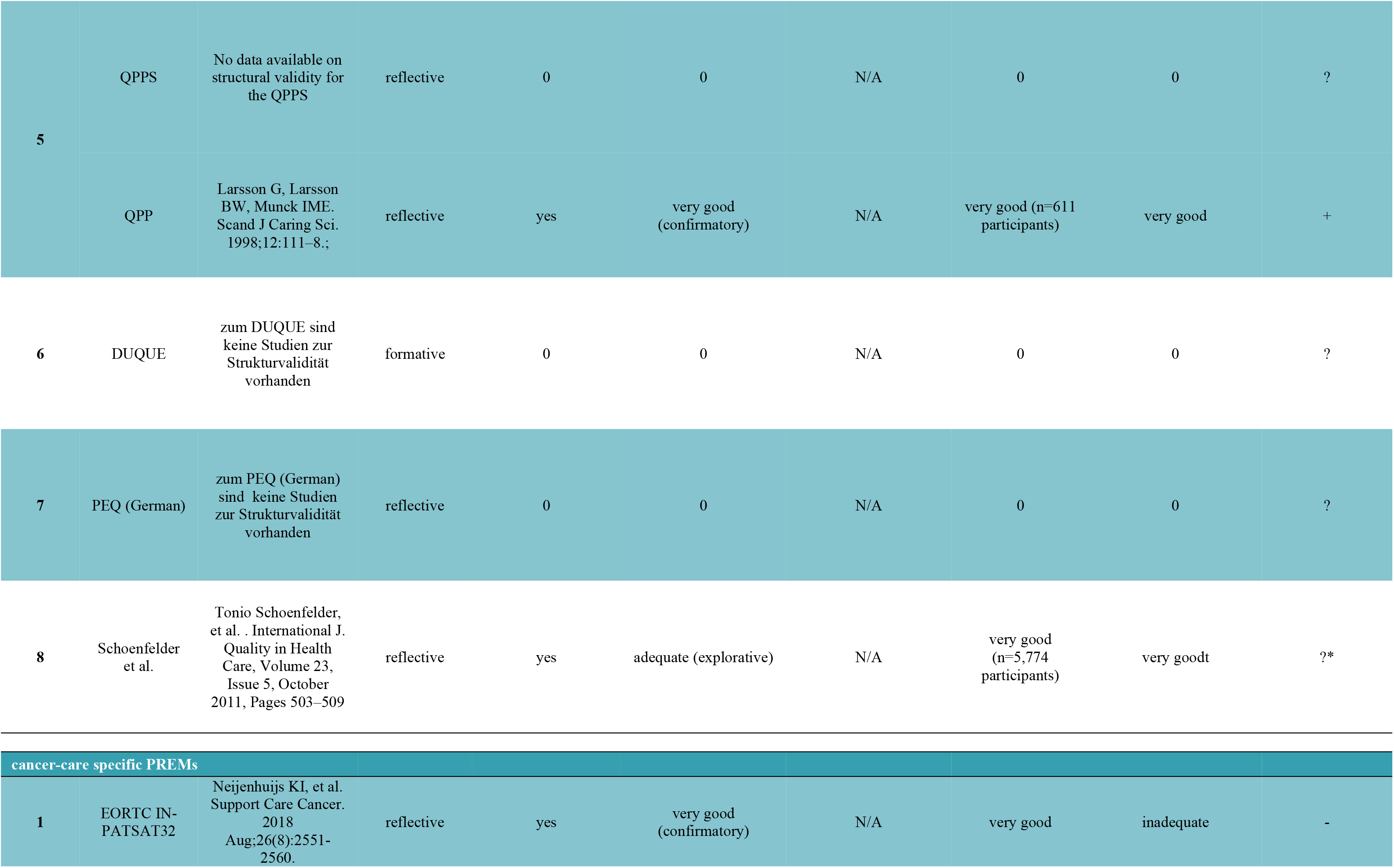
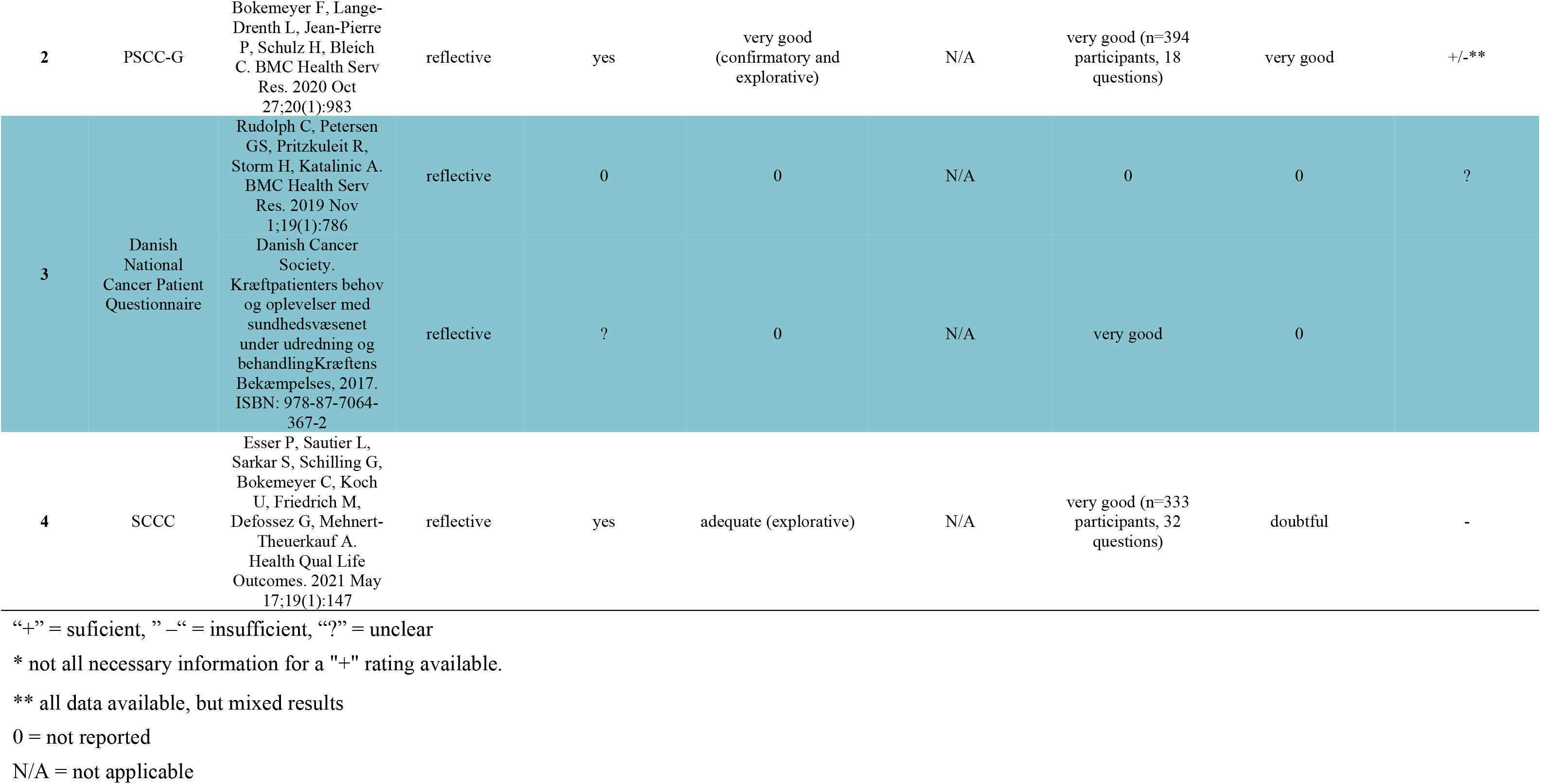
Overview of the analyses results on structural validity of German-language PREMs.

### Internal consistency

Table 6 shows the results of internal consistency studies of German-language PREMs. Cronbach’s Alpha for the NORPEQ (containing only one scale) is 0.85 [20]. A single validation study is available for the PPE-15 [16]. It is unclear whether the consistency statistics contained in this study are calculations of Cronbach’s Alpha or not. Furthermore, the consistency analyses are not available for the various PPE-15 subscales, which are partly dichotomous, partly continuous. Consequently, the PPE-15 received an “insufficient” rating for study design. Similarly, internal consistency analyses (Cronbach’s Alpha) of the PEQ refer to the entire questionnaire and not to the various subscales [25]. There are no internal consistency studies for the PEACS questionnaire. Keller et al. reported data on internal consistency for all 7 subscales of the HCAHPS [19]. With exception of the subscale “*Medicine Communication*“ (Cronbach’s Alpha 0.66) all HCAHPS subscales show a Cronbach’s Alpha ≥0.70. Analysis of internal consistency is available for all subscales of the QPPS in the study by Larsson et al. [18]. Results are mixed as some subscales exhibit Cronbach’s alpha ≥0.70 and some <0.70. No data on internal consistency could be found for the DUQUE and the *Danish National Cancer Patient Questionnaire.* For the PREM by Schoenfelder et al. internal consistency results are available for both subscales of the questionnaire (*received service* and *medical aspects).* Both of which have a Cronbach’s Alpha ≥0.70.

**Table 6.**
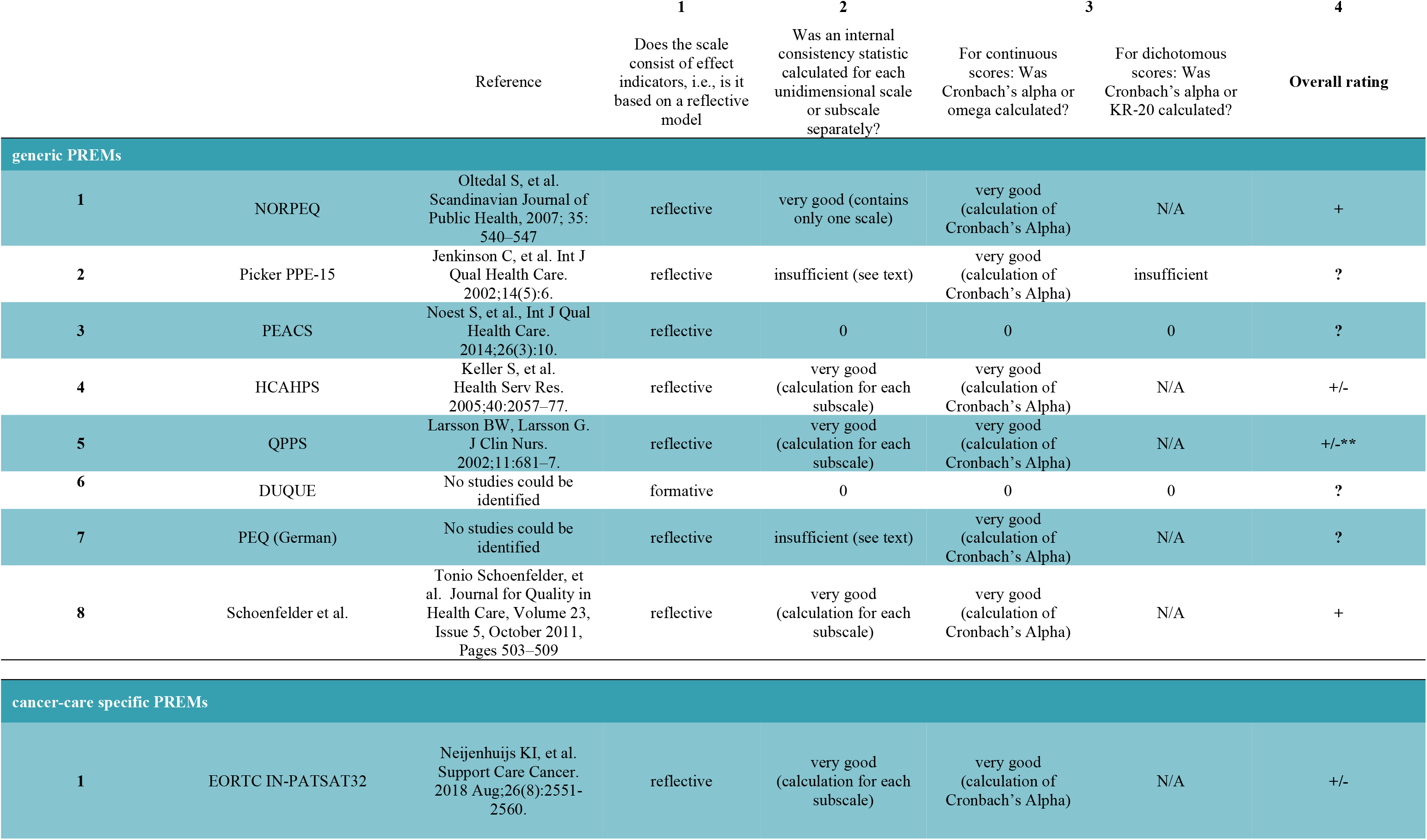
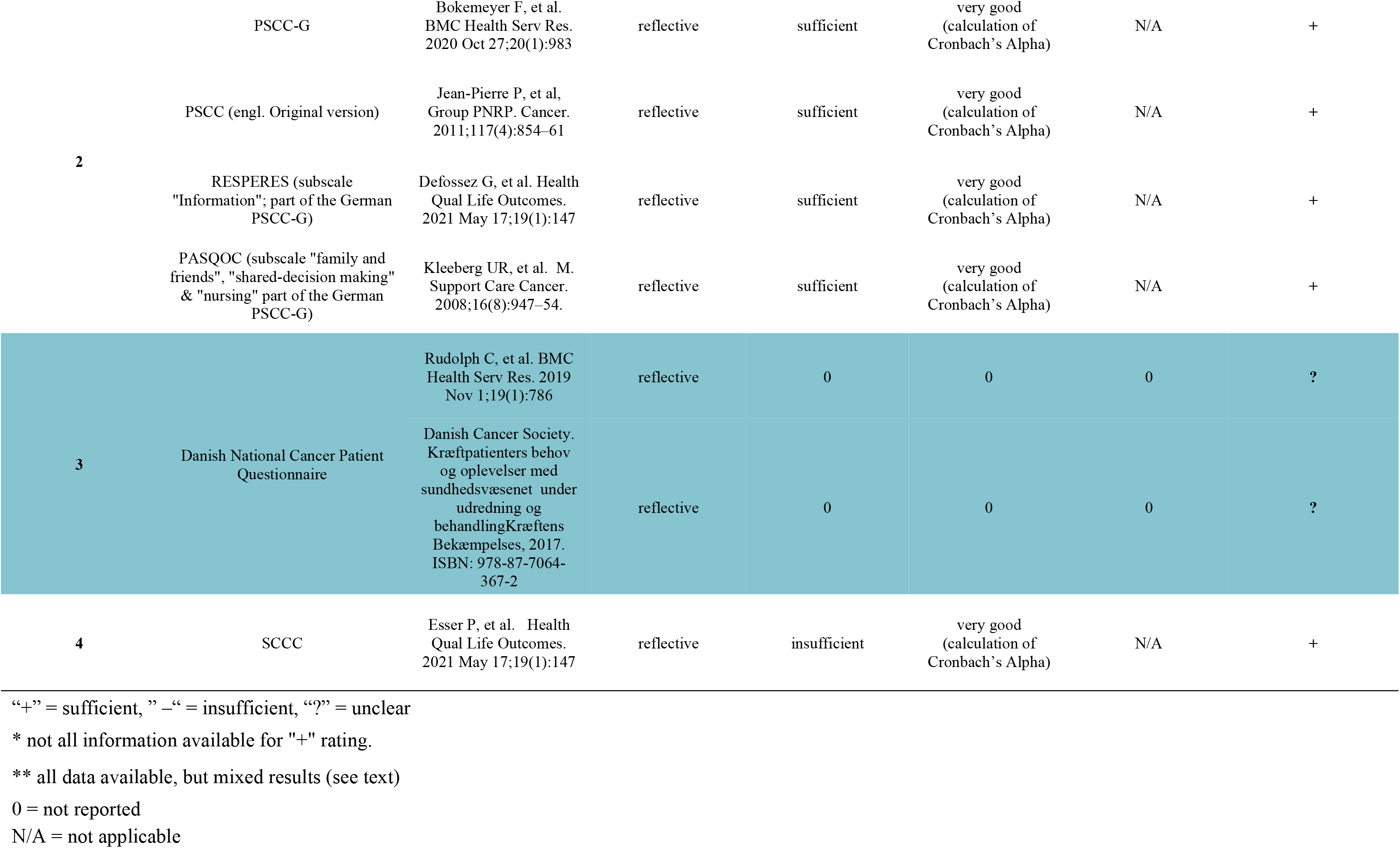
Overview of internal consistency studies of German-language PREMs. Rating according to COSMIN guidelines.

The internal consistency of the EORTC IN-PATSAT32 has been analyzed in a recent metaanalysis [29]. According to this metaanalysis, 5 studies are available on the internal consistency of the IN-PATSAT32, of which 3, however, are of poor methodological quality. Analyses are available for all subscales of the questionnaire, with Cronbach’s Alpha ≥0.70 across all scales, except for the subscale *hospital access* (Cronbach’s Alpha <0.70). The overall Cronbach’s Alpha for the entire PSCC-G is reported to be 0.92 [22]. As the German version of the PSCC (PSCC-G) consists of the German translation of the English PSCC [30] as well as of translations of parts of the French REPERES (subscale „Information“) [31] and German PASQOC [32] (subscales *family and friends*, *shared-decision making* and *nursing staff*), the original questionnaires can be analyzed for internal consistency as well. For all subscales Cronbach’s Alpha is reported to be ≥0.70.

### Measurement invariance / Cross-cultural validity

“Cross-cultural validity refers to the degree to which the performance of the items on a translated or culturally adapted instrument are an adequate reflection of the performance of the items of the original version of the instrument” [11]. It is particularly important for PREM translations into German. Results are depicted in Table 7. No data on cross-cultural validity could be found for the DUQUE questionnaire. A cross-cultural validation study for the NORPEQ is available is available for several Scandinavian countries, but not for German. The PPE-15 underwent validation in 5 countries including German speaking countries (Switzerland and Germany) [16]. The patient cohorts were comparable in respect to sex and age, but not in respect to indications (elective vs. emergency). No further details about the patient cohorts were collected. Furthermore, no adequate study could be identified that compares the English original questionnaire with the German translation [16]. Such a study exists for the Spanish language [33]. This study shows that an expansion of the questionnaire from 15 to 33 questions (PPE-33) is necessary to preserve psychometric properties [33]. There is a cross-cultural validation study for the HCAHPS PREM [34]. In this study it is unclear whether patient cohorts were comparable. Furthermore, only 10 German-speaking patients were included [34]. In addition, the statistical methods used are considered to be inadequate according to current COSMIN guidelines. The QPPS has been used in two German studies without prior validation of the German translation [28, 35]. Validation of the EORTC IN-PATSAT32 occurred in an international study in which 34 German-speaking patients were included [36]. However, cross-cultural validity was not analyzed in this study. An addition, the EORTC IN-PATSAT32 metaanalysis by Neijenhuijs et al. found no studies on cross-cultural validation of the questionnaire in German [29]. Similarly, no cross-cultural validation in German could be found for the Danish *National Cancer Questionnaire* [37]. Rudolph et al. use the Danish *National Cancer Questionnaire* in their study but without previous validation [21]. The PSCC-G has been validated in German in a study by Bokemeyer et al. with a sufficient number of patients and adequate statistical methods [22]. The remaining PREMs (PEACS, PEQ, Schoenfelder et al, SCCC) have been developed in German including patients from different medical specialities.

**Table 7.**
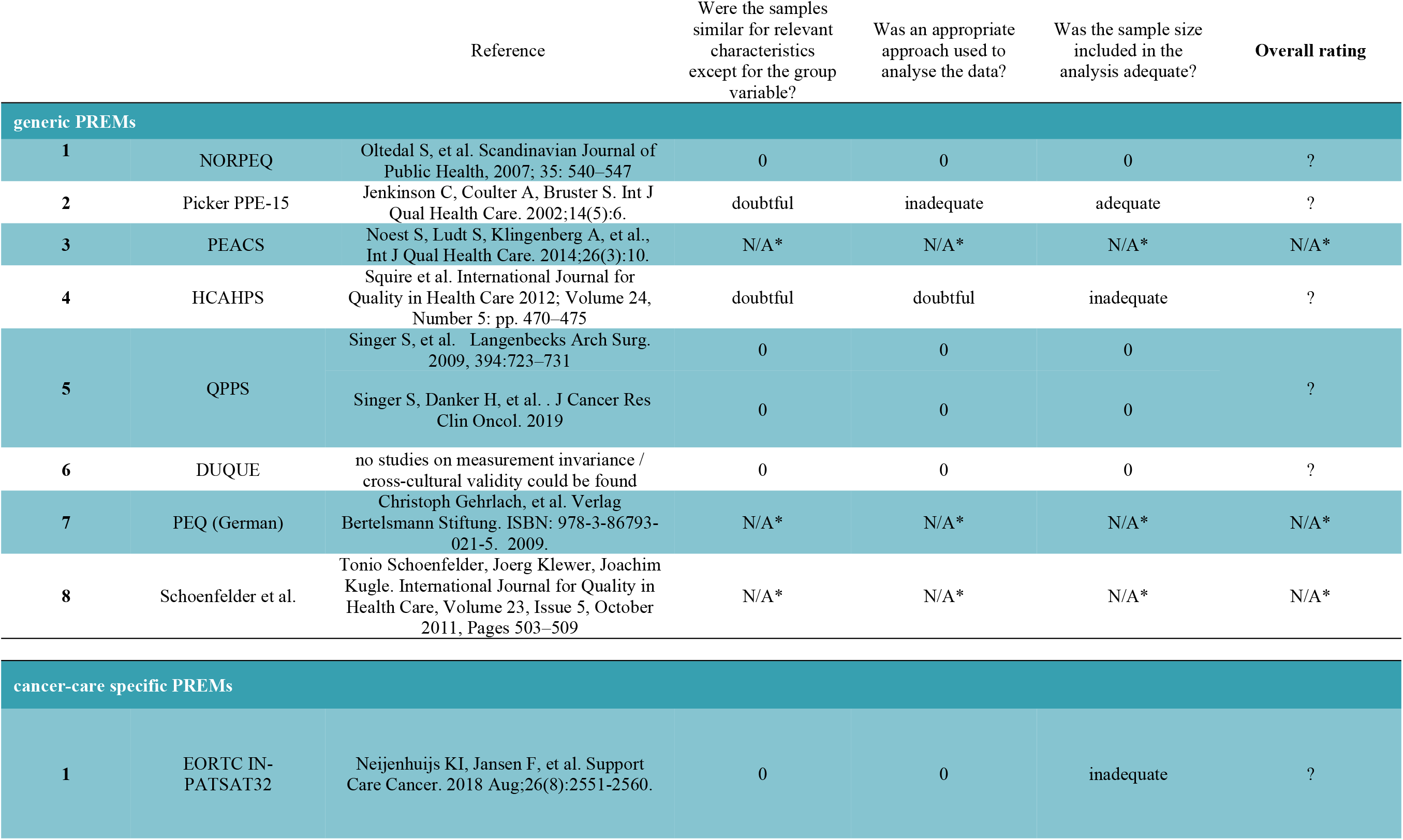
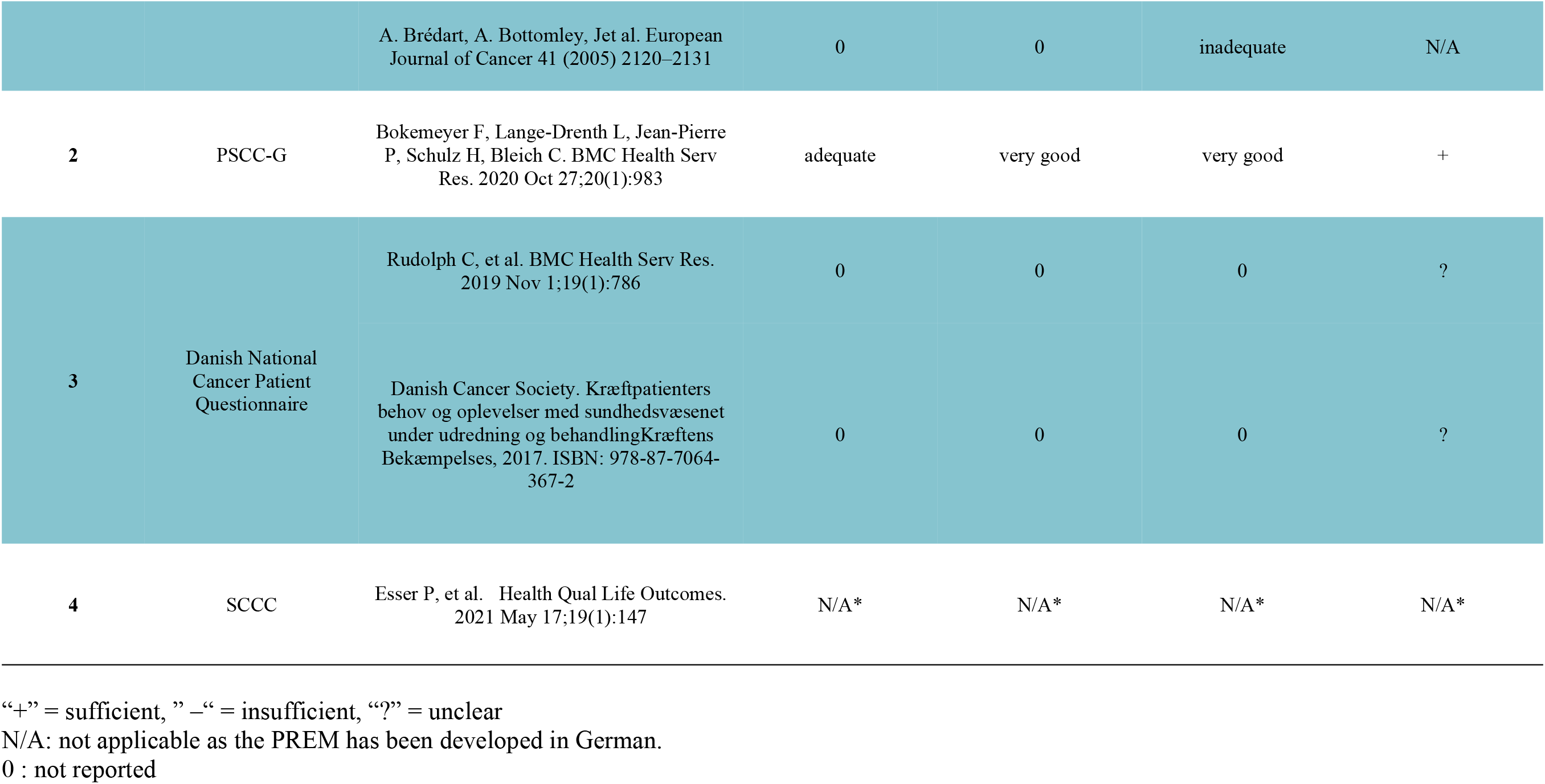
Results of cross-cultural validity analyses of PREMs into German. Rating according to COSMIN guidelines.

### Reliability

An overview of the results of the reliability analyses can be found in table 8. A study measured test-retest reliability of the NORPEQ [20] for which 68 of 244 patients were resent the questionnaire within 5-6 days. The intraclass correlation index (ICC) was between 0.45 (*nurse professional skills*) and 0,83 (*doctors understandable*). Four of the eight subscales exhibited an ICC <0.70. The test-retest reliability for the total score was 0.88 [20]. Consequently, the overall reliability rating for the NORPEQ was +/-. Noest et al. analyzed reliability for the PEACS questionnaire [24]. Test-retest reliability was measured via weighted kappa. With exception of the subscale institutional treatment and transition (weighted kappa 0,671), the weighted kappa was ≥0.70 for all subscales. Thus, overall rating was +. The study by Keller et al. analyzed hospital-level reliability [19]. No data on test-retest reliability could be identified for the HCAHPS. Hospital-level reliability assumes that recurrent measurements (retesting) of patients in the same hospital should be more similar than recurrent measurements in another hospital. In the study by Keller et al. 300 patients from different hospitals were asked to fill out the HCAHPS. For six of the seven subscales the ICC value was ≥0.70. The only exception was *medicine communication* (ICC 0,66).

**Table 8.**
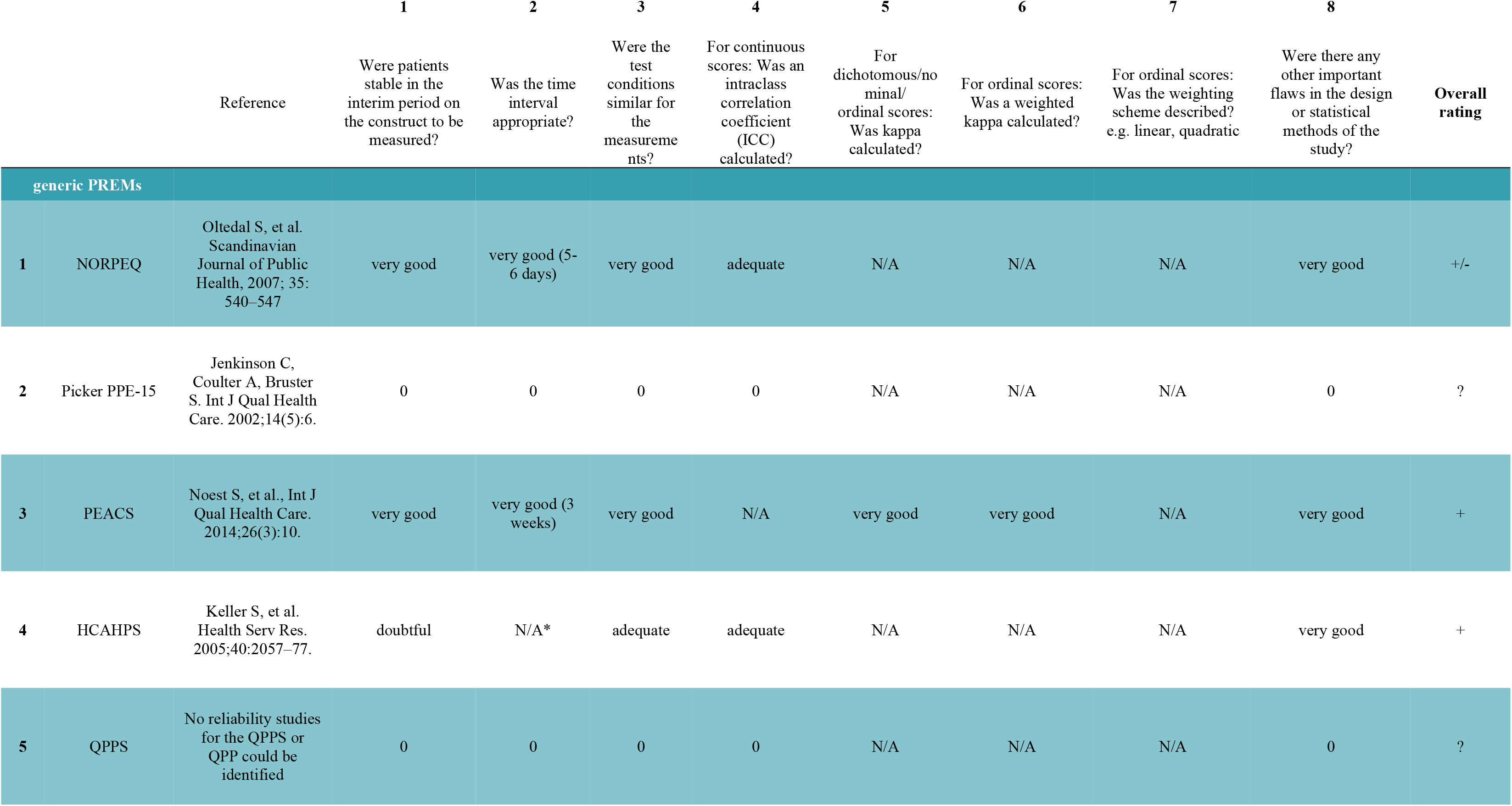
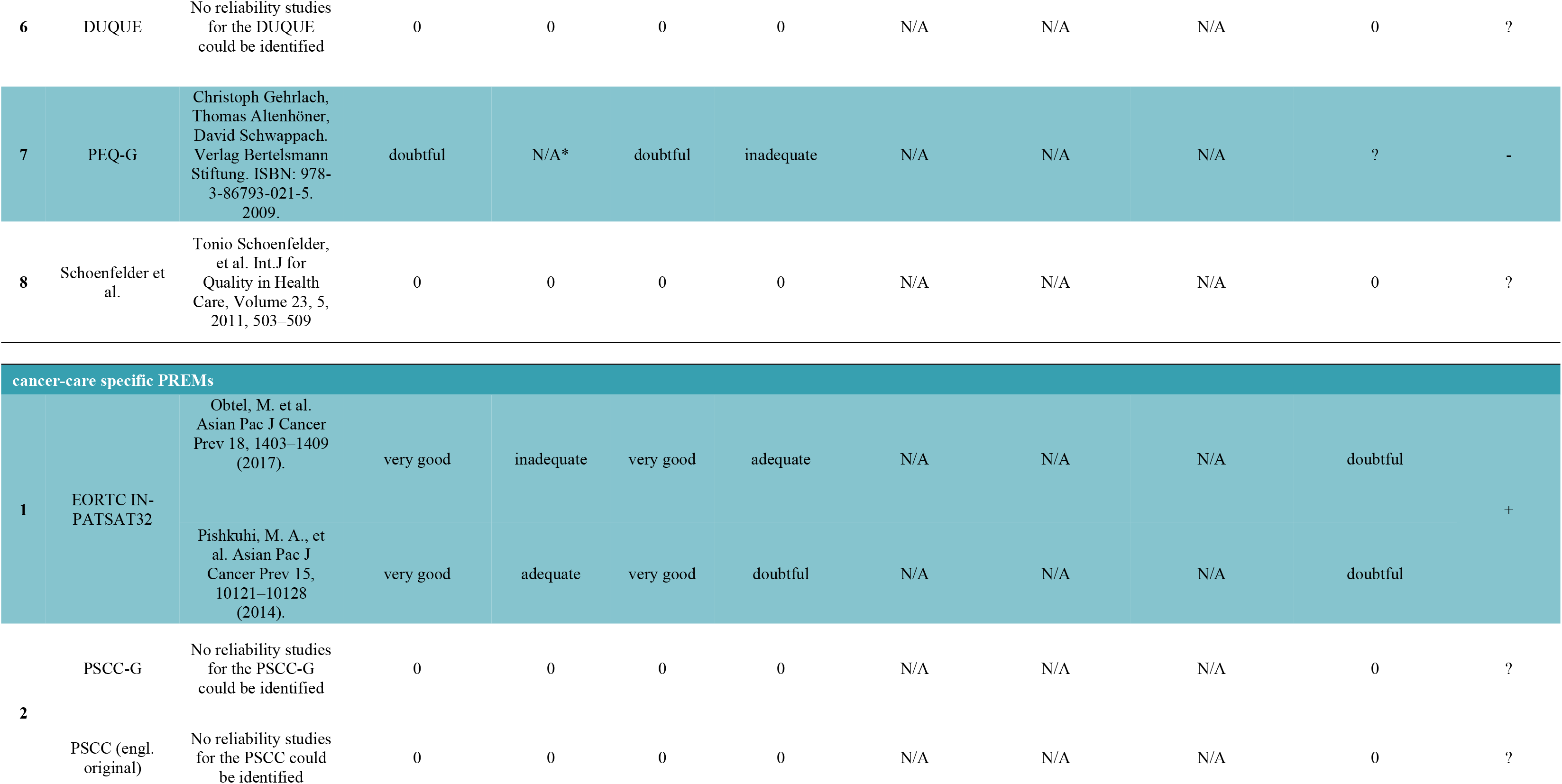
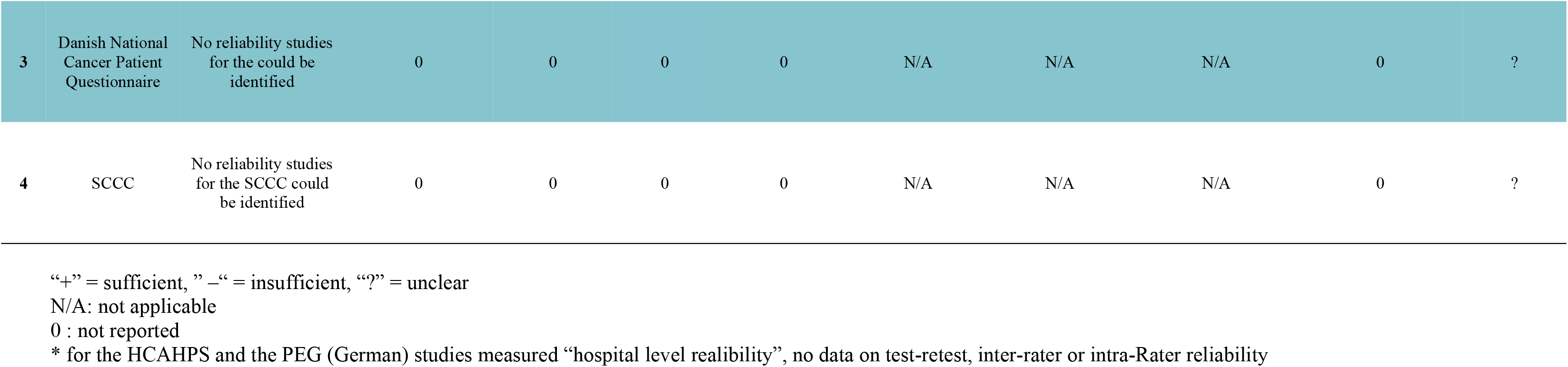
Results of reliability analyses of German-language PREMs. Rating according to COSMIN guidelines.

The overall rating for the HCAHPS was +. A similar hospital-level reliability analysis can be found for the PEQ in the study by Gehrlach et al [25]. Logistic regression analyses were used to find out whether the questionnaire can distinguish between patient cohorts from different hospitals. However, no specific results are reported except for “…none of the instruments showed significant results”. Further reliability data for the PEQ could not be identified. No reliability study could be identified for the PPE-15, QPPS or the QPP, DUQUE, PSCC-G, the Danish National Cancer Patient Questionnaire or the SCCC.

For the cancer-care specific PREM EORTC IN-PATSAT32 two studies investigate test-retest reliability [38, 39]. Appreciation of reliability in these two studies has already been done by Neijenhuijs et al. [29]. ICC was ≥0.70 for all subscales of the EORTC IN-PATSAT32 in the study by Pishkuhi et al. In the study by Obtel et al. all scales except *doctor availability* (correlation coefficient 0.64) and *overall satisfaction* (correlation coefficient 0.67) showed a correlation coefficient ≥0.70. Consequently, overall reliability rating was +. However, both studies showed methodological weaknesses as the time interval between test and retest was too short (30 minutes) [39] or it was unclear, which type of correlation coefficient has been used [38].

### Analysis of measurement error

An overview of the results for the analysis of measurement error can be found in Table 9. Measurement error could not be analyzed as *no minimal important change* has been defined for any of the German PREMs so far. Therefore, all PREMs received “?” rating. Only for HCAHPS there has been a calculation of the standard error of Measurement (SEM). For the EORTC IN-PATSAT32 the SEM and SDC can be calculated from the studies by Obtel et al. [39] and Pishkuhi et al. [38] as has been shown by Neijenhuijs et al. [29]. However, as no *minimal important change* has been defined for the EORTC IN-PATSAT32 an overall rating of the measurement error is not possible.

**Table 9.**
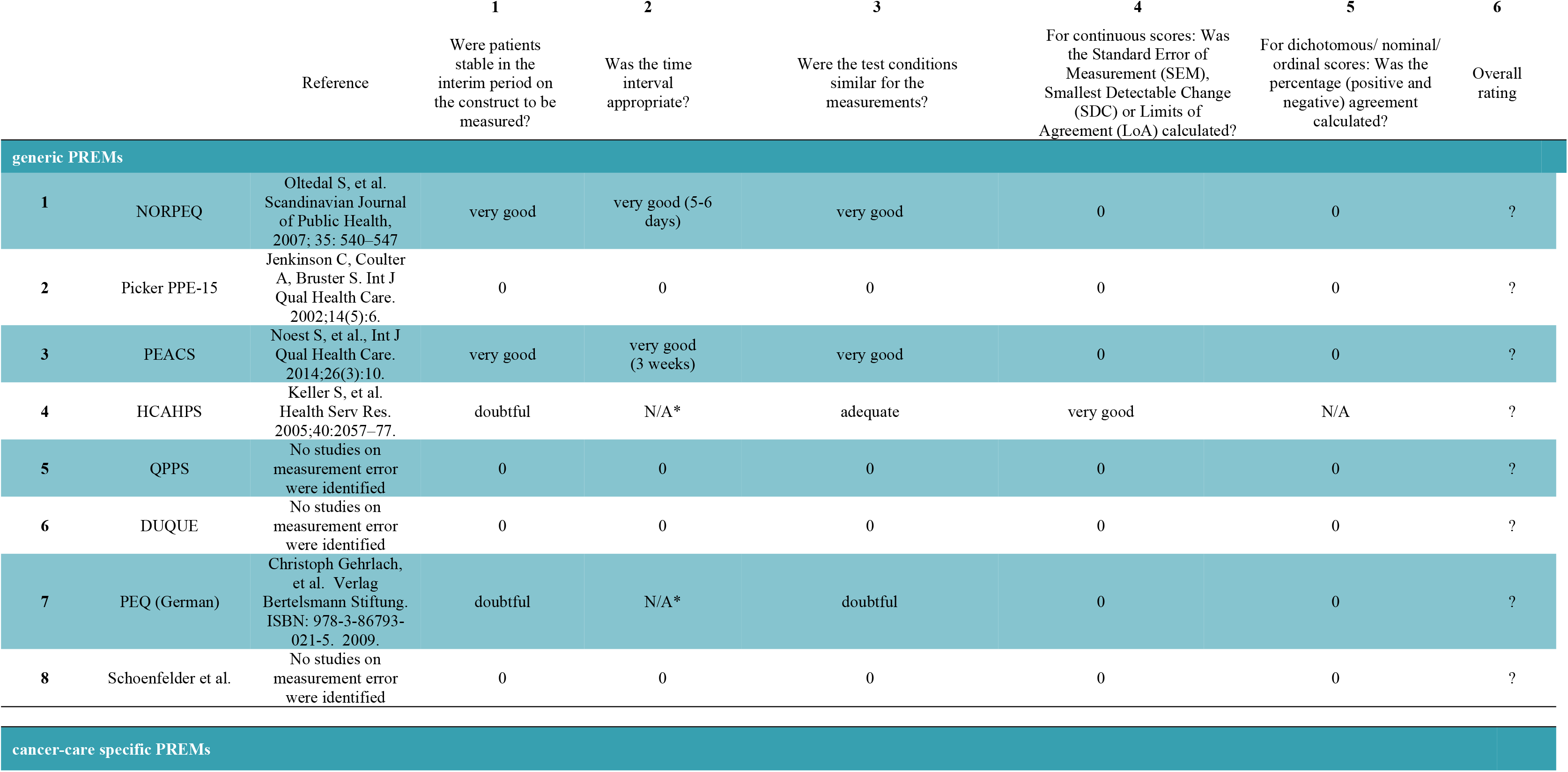
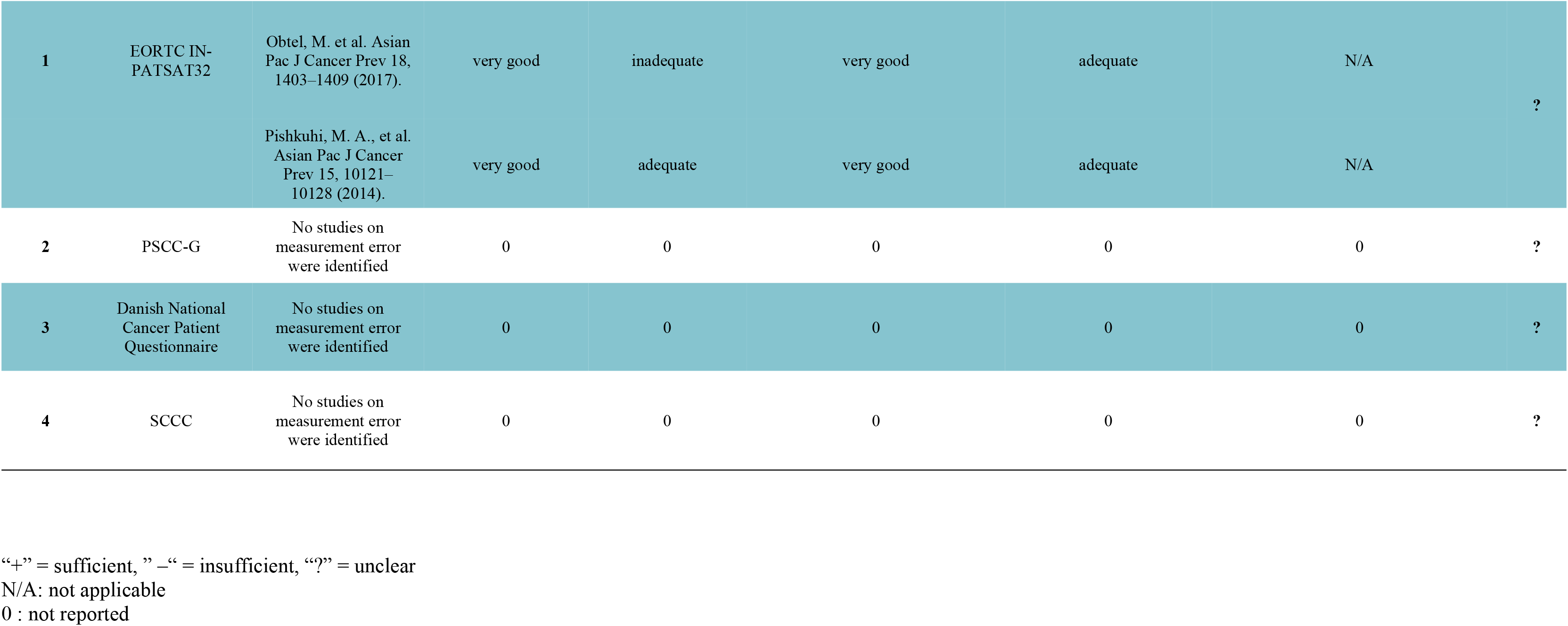
Results of analyses for measurement error of German-language PREMs. Rating according to COSMIN guidelines.

|  |  | Reference | 1<br>Were patients<br>stable in the<br>interim period on<br>the construct to be<br>measured? | 2<br>Was the time<br>interval<br>appropriate? | 3<br>Were the test conditions<br>similar for the<br>measurements? | 4<br>For continuous scores: Was<br>the Standard Error of<br>Measurement (SEM),<br>Smallest Detectable Change<br>(SDC) or Limits of<br>Agreement (LoA) calculated? | 5<br>For dichotomous/ nominal/<br>ordinal scores: Was the<br>percentage (positive and<br>negative) agreement<br>calculated? | 6<br>Overall<br>rating |
| --- | --- | --- | --- | --- | --- | --- | --- | --- |
| **generic PREMs** |  |  |  |  |  |  |  |  |
| 1 | NORPEQ | Oltedal S, et al.<br>Scandinavian Journal<br>of Public Health,<br>2007; 35: 540–547 | very good | very good (5-6<br>days) | very good | 0 | 0 | ? |
| 2 | Picker PPE-15 | Jenkinson C, Coulter<br>A, Bruster S. Int J<br>Qual Health Care.<br>2002;14(5):6. | 0 | 0 | 0 | 0 | 0 | ? |
| 3 | PEACS | Noest S, et al., Int J<br>Qual Health Care.<br>2014;26(3):10. | very good | very good<br>(3 weeks) | very good | 0 | 0 | ? |
| 4 | HCAHPS | Keller S, et al.<br>Health Serv Res.<br>2005;40:2057–77. | doubtful | N/A* | adequate | very good | N/A | ? |
| 5 | QPPS | No studies on<br>measurement error<br>were identified | 0 | 0 | 0 | 0 | 0 | ? |
| 6 | DUQUE | No studies on<br>measurement error<br>were identified | 0 | 0 | 0 | 0 | 0 | ? |
| 7 | PEQ (German) | Christoph Gehrlach,<br>et al. Verlag<br>Bertelsmann Stiftung.<br>ISBN: 978-3-86793-021-5. 2009. | doubtful | N/A* | doubtful | 0 | 0 | ? |
| 8 | Schoenfelder et al. | No studies on<br>measurement error<br>were identified | 0 | 0 | 0 | 0 | 0 | ? |
| **cancer-care specific PREMs** |  |  |  |  |  |  |  |  |
| 1 | EORTC IN-PATSAT32 | Obtel, M. et al. Asian Pac J Cancer Prev 18, 1403–1409 (2017). | very good | inadequate | very good | adequate | N/A | ? |
|  |  | Pishkuhi, M. A., et al. Asian Pac J Cancer Prev 15, 10121–10128 (2014). | very good | adequate | very good | adequate | N/A |  |
| 2 | PSCC-G | No studies on measurement error were identified | 0 | 0 | 0 | 0 | 0 | ? |
| 3 | Danish National Cancer Patient Questionnaire | No studies on measurement error were identified | 0 | 0 | 0 | 0 | 0 | ? |
| 4 | SCCC | No studies on measurement error were identified | 0 | 0 | 0 | 0 | 0 | ? |

“+” = sufficient, ” –“ = insufficient, “?” = unclear
N/A: not applicable
0 : not reported

### Analysis of criterion validity

As no gold standard for the measurement of patient-centeredness has yet been defined, overall criterion validity cannot be analyzed for PREMs. However, for some PREM subscales, gold standards for measurement are available. Consequently, criterion validity for these subscales can be analyzed. No data on criterion validity was found for NORPEQ, PPE-15, HCAHPS, QPPS (or QPP), DUQUE, Schoenfelder et al., EORTC IN-PATSAT-32, PSCC, SCC and the Danish National Cancer Patient Questionnaire.

The PEACS subscales *shared decision making* and *information at discharge* were compared to the two validated and established German questionnaires *Shared Decision Making Questionnaire* (SDM-Q-9) [40] and the *Care-Transition-Measurement* Questionnaire (CTM) [41], respectively. Both subscales showed a very high (SDM-Q-9; r = 0.814, p < 0.001) or high correlation (CTM-3; r = 0.511, p<0.001) with the respective gold standard [24].

The PEQ was correlated to the Cologne Patient Questionnaire *(Kölner Patientenfragebogen, KPF)*. For the PEQ subscales “physicians” and “nursing” there was a high correlation >0.70 with the KPF. Only a weak correlation was found for the subscale “management” (between −0,28 and −0,46) [25].

### Analysis of hypothesis testing for construct validity

Hypothesis testing for construct validity describes the degree to which a PREM results is consistent with an *a priori* hypothesis. The hypothesis to be tested can either be a comparison of PREM results between two clinically defined patient groups (*known groups validity)* or PREM subscales can be compared to another known measurement tool *(convergent validity)*. No data on hypothesis testing for construct validity could be found for: PEACS, DUQUE, PEQ-G, *Danish National Cancer Patient Questionnaire.* Results for all other German PREMs can be found in Table 10. Most hypotheses are tested positive, i.e., results confirm the a priori formulated hypothesis. However, all PREMs, except for the PSCC-G, also show negative results. For the PSCC-G only positive test results could be found.

**Table 10.**
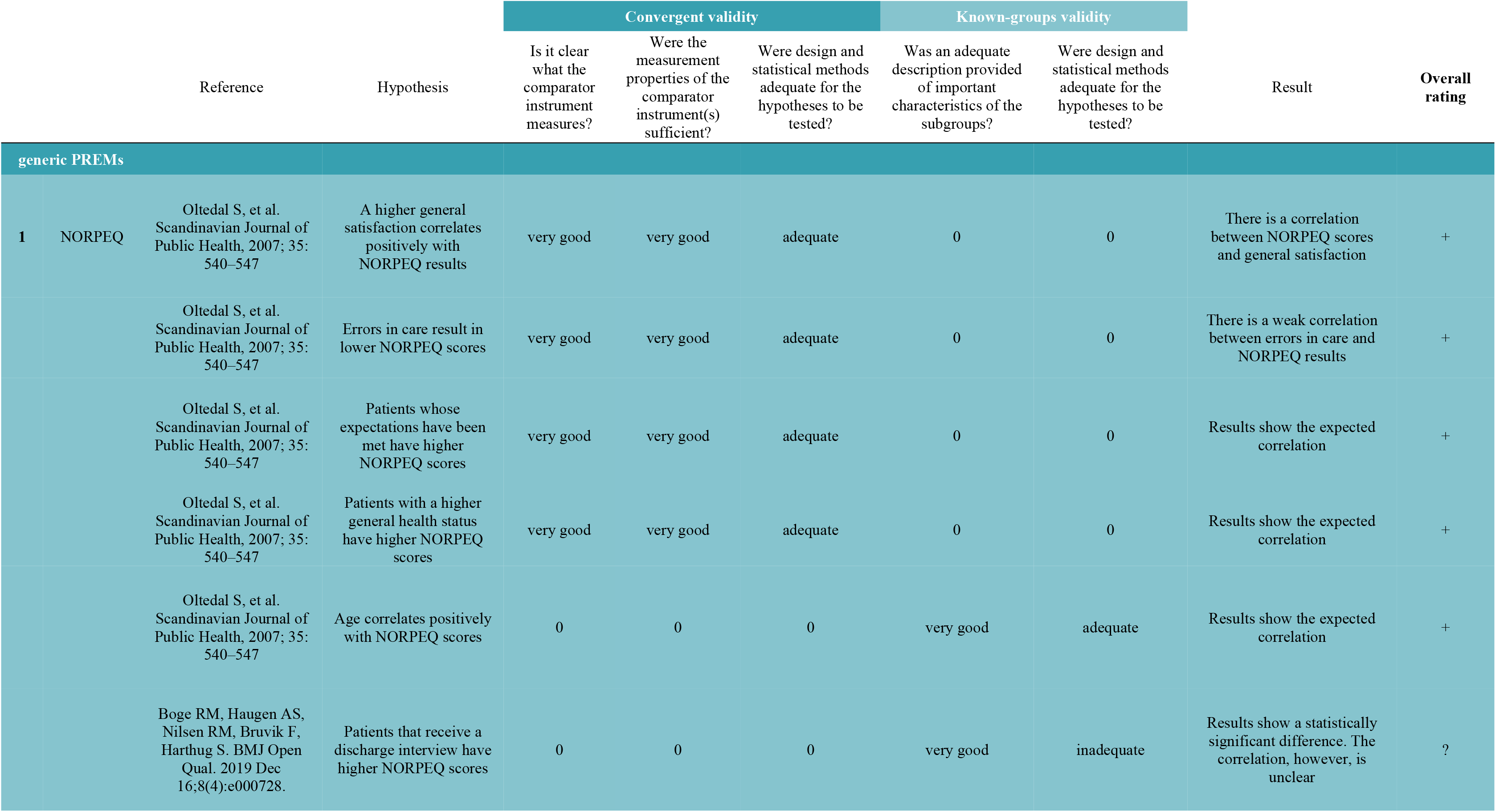
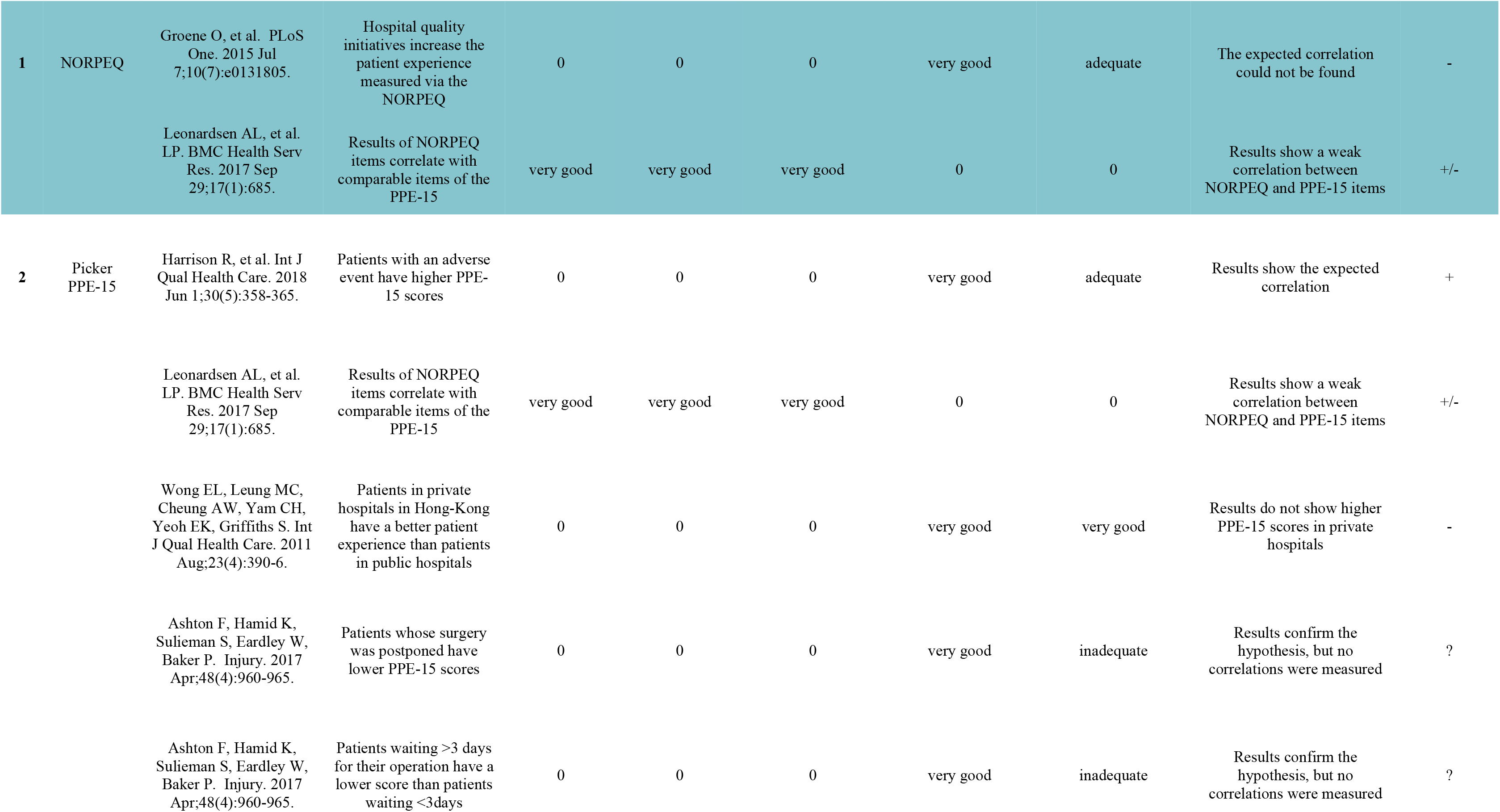
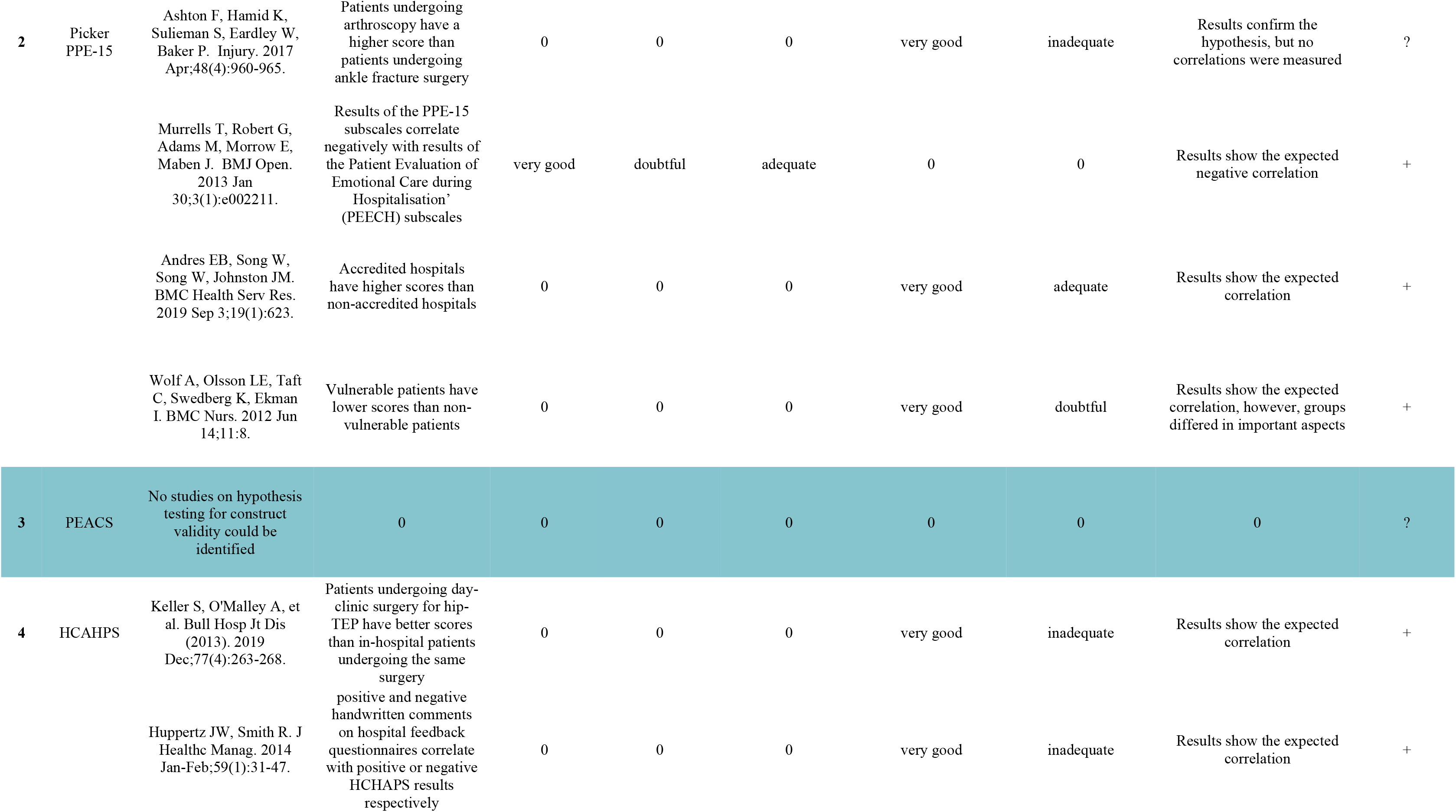
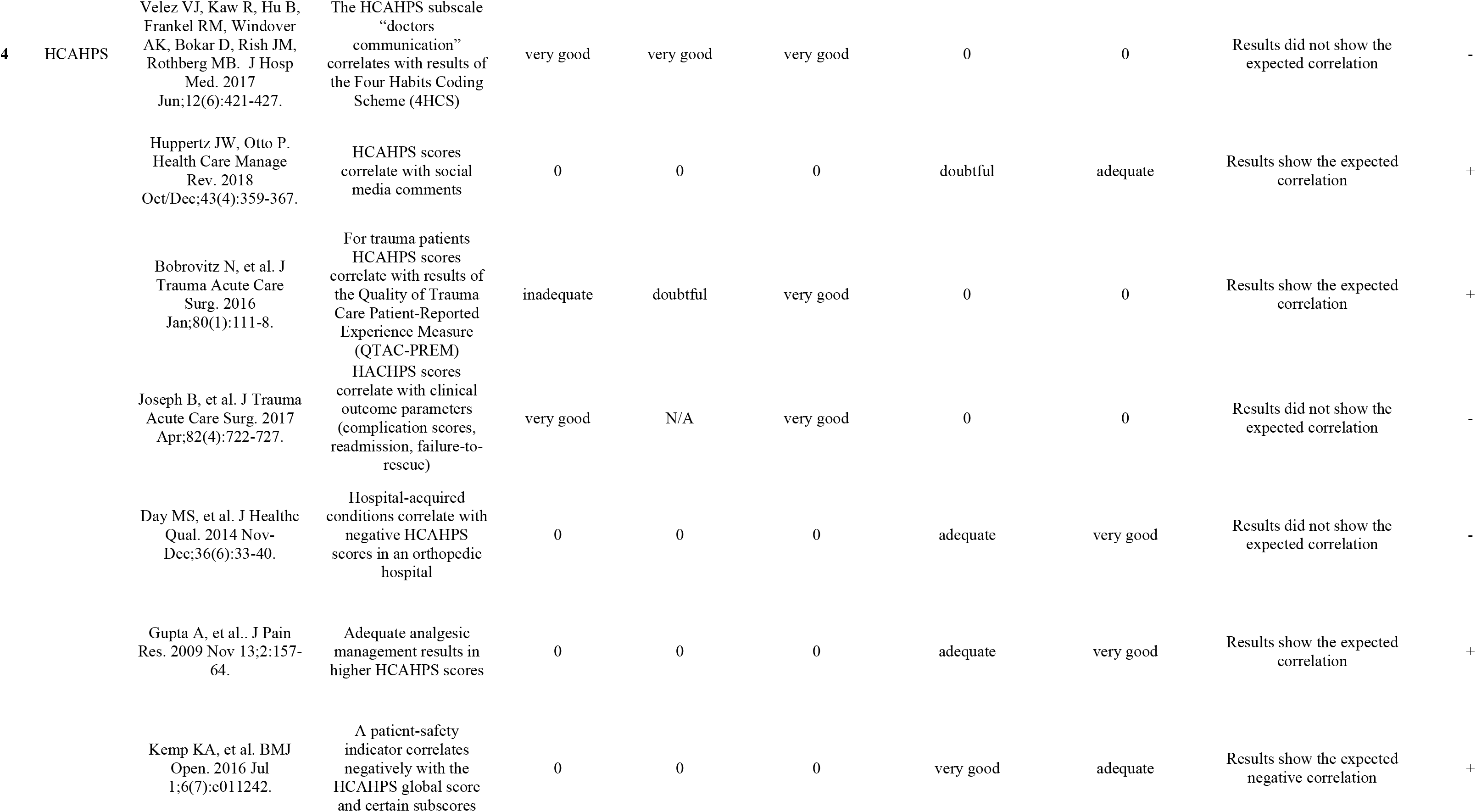
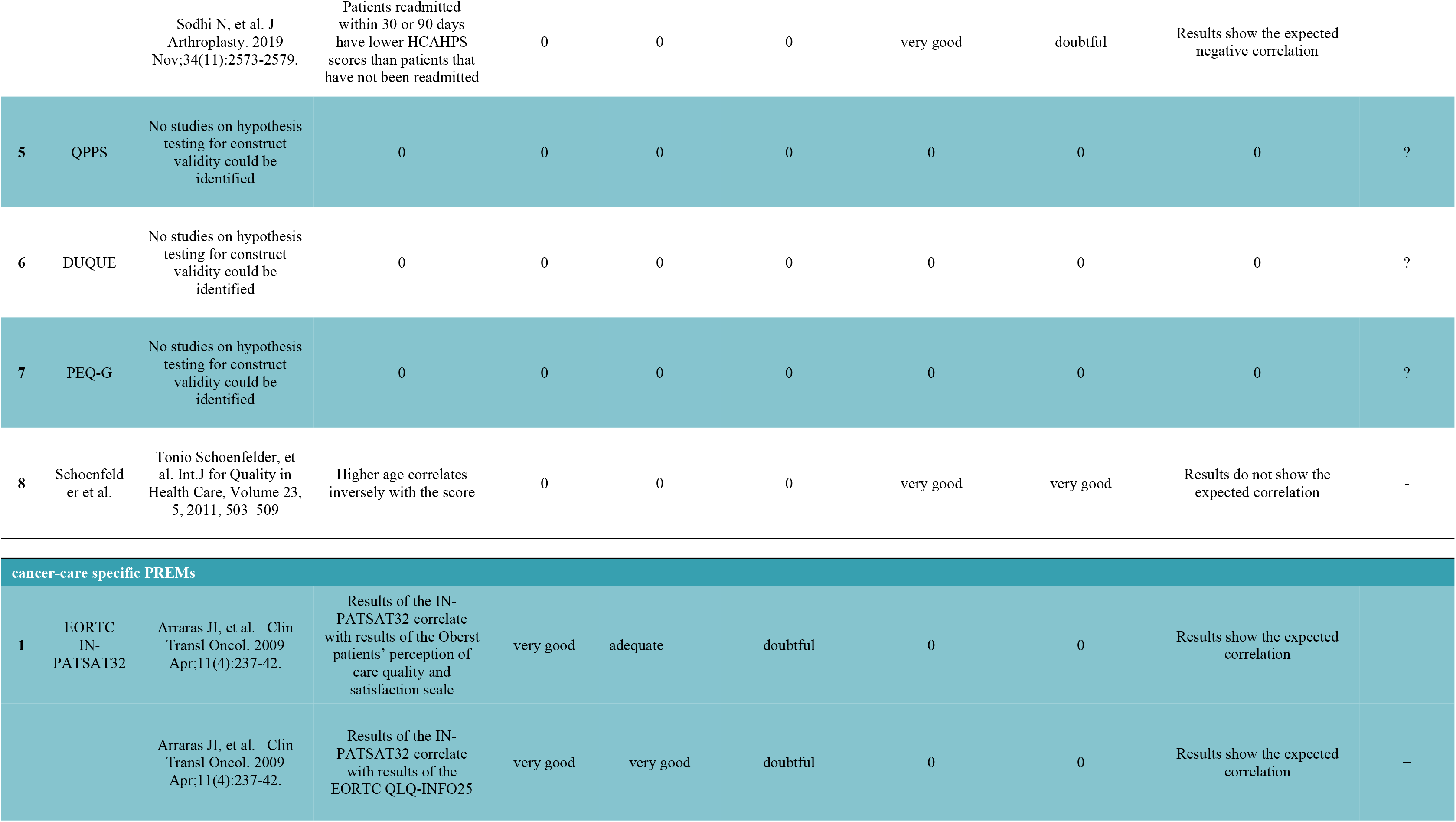
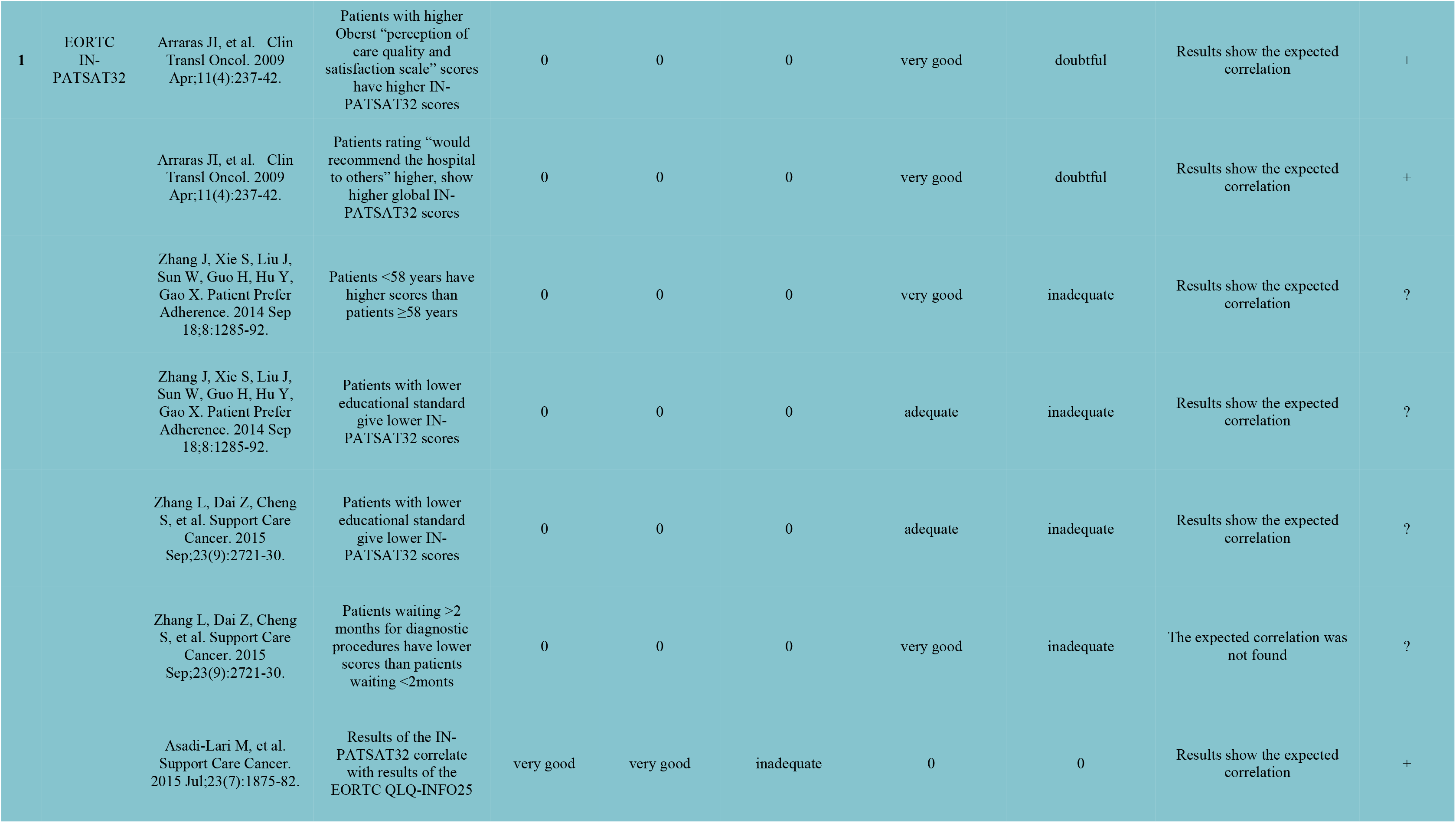
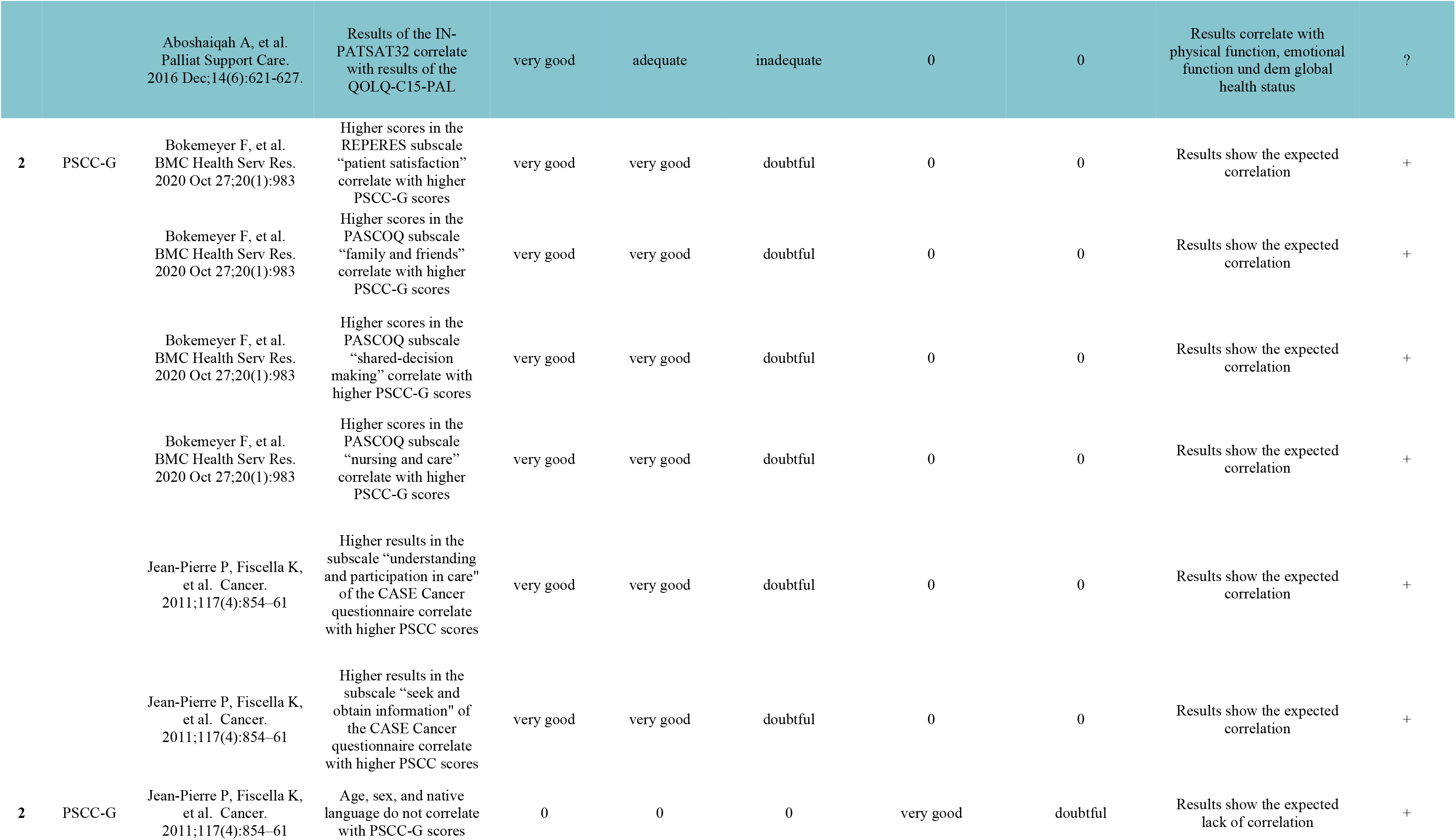
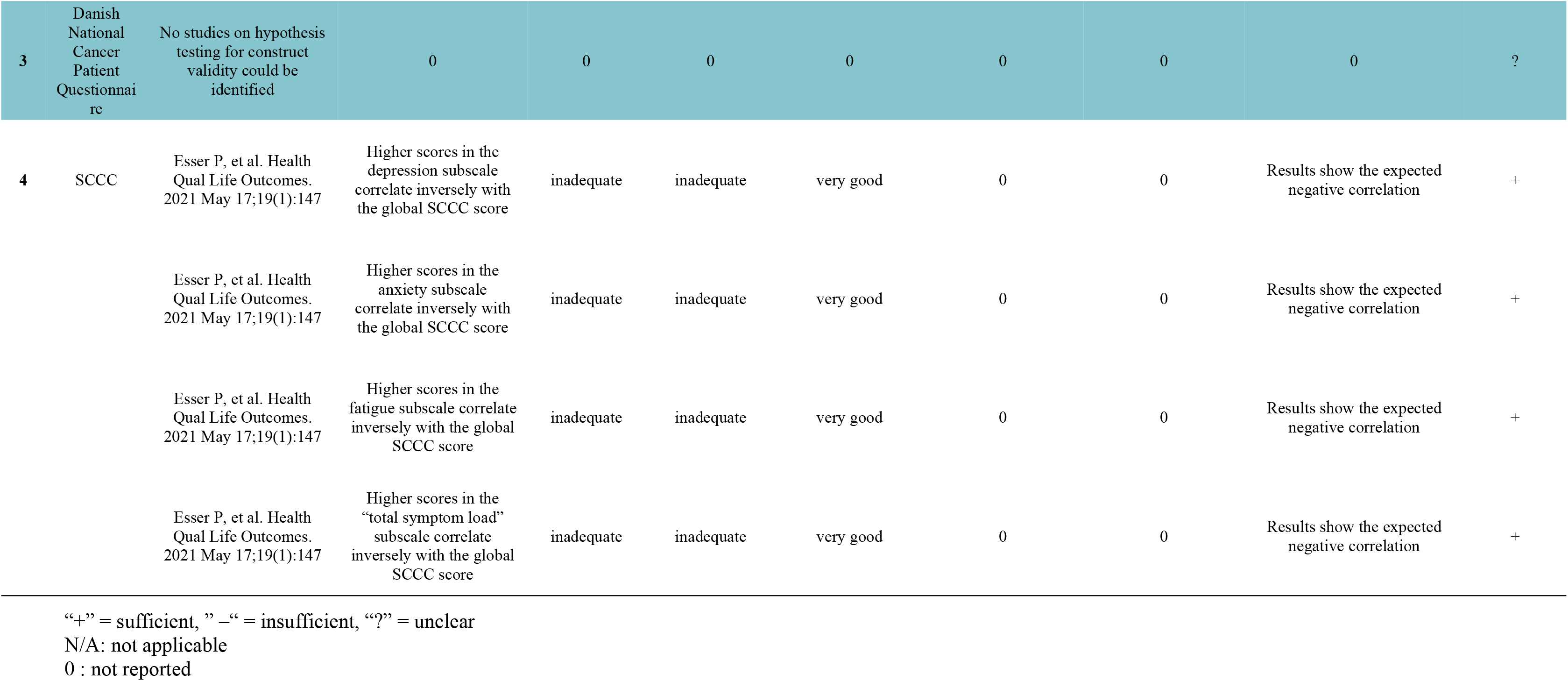
Results of hypothesis testing for construct validity of German-language PREMs. Rating according to COSMIN guidelines.

## Discussion

In the current study numerous German language PREMs could be identified that were not contained in previous publications [8, 42]. This was due to publications in recent years as well as due to the difficulty in identifying PREMs by database searches alone. Many German PREMs were found by hand-searches. The current study uses for the first time the current COSMIN guidelines for the assessment of PREMs [15]. Furthermore, by using a comprehensive model of PC covering all dimensions of PC (S2 Table) a thorough analysis of content validity of PREMs was possible for the first time. The study uncovers a deficit in German language PREMs with adequate reliability and validity. The analysis of content validity shows that there is no “perfect” PREM, but rather several available tools each with its strengths and weaknesses. A context-specific application of German PREMs is mandatory and several recommendations can be made.

### Recommendations

Two PREMs cannot be recommended currently for use in German because of a lack of validation of psychometric properties: the DUQUE questionnaire [27], as well as the German translation of the *Danish National Cancer Patient Questionnaire* used by Rudolph et al. [21]. Depending on intended use, one of the remaining ten PREMs can be selected. Fig 2 shows a schematic representation of the remaining PREMs within their intended area of use. The figure can facilitate selection. In a next step, the results of this systematic review can be used to select a PREM with sufficient psychometric properties (Table 5-10) and the necessary content (Table 3 and 4). For example, for cancer healthcare-specific PREMs the PSCC-G has significant better psychometric properties than the SCC with its insufficient structural validity and lack of assessment in many psychometric domains. Furthermore, when selecting a PREM the intended area of application should to be considered [8]: is it intended as a reflection instrument for patients or rather as a provider-specific evaluation instrument for internal use or as a benchmarking instrument to compare different providers? In each case different content dimensions (Table 3) and length of questionnaires (Table 2) are of interest.

**Fig 2.**
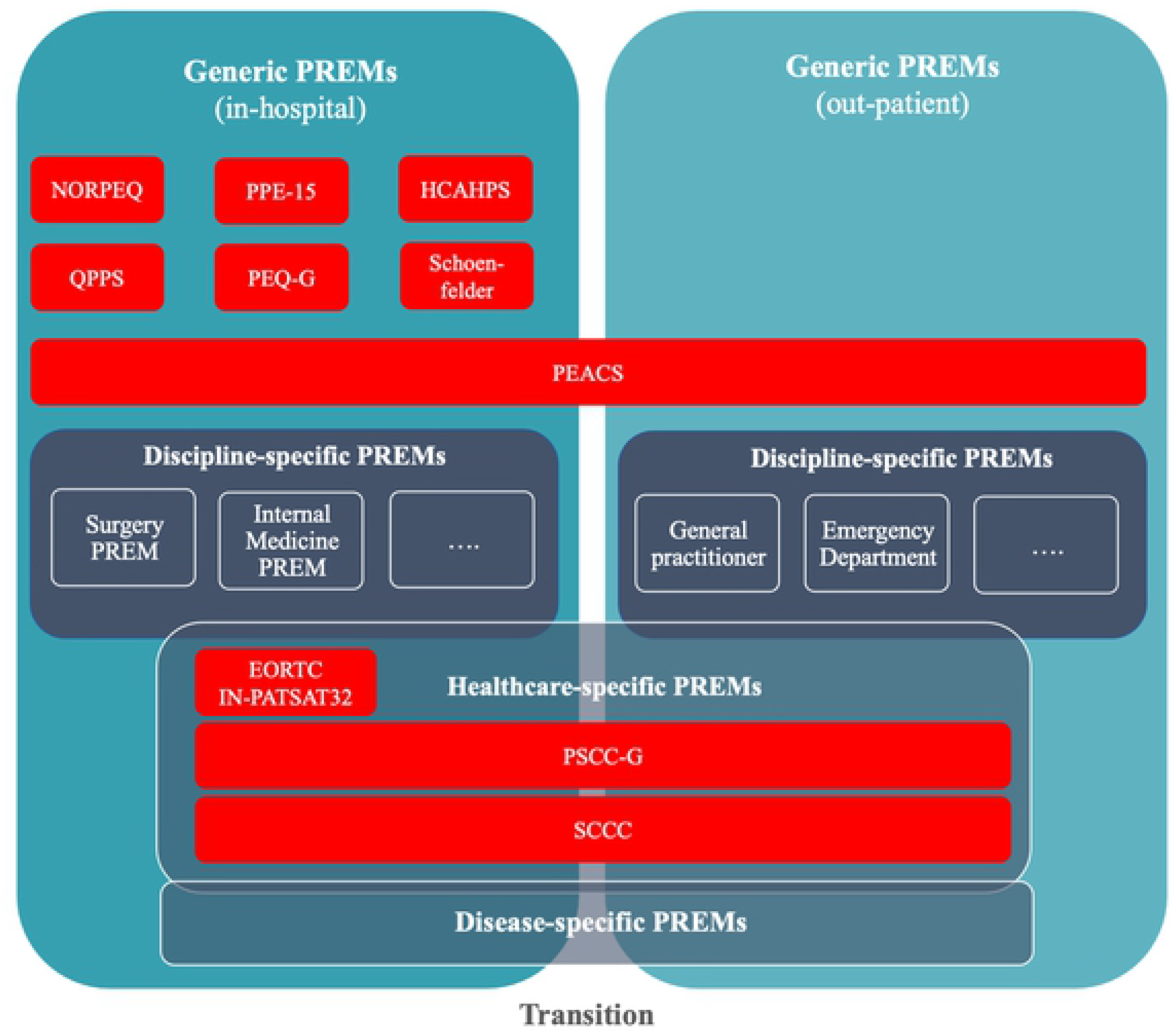
German language Patient-Reported Experience Measures (PREMs). Overview of German PREMs in their intended category of use

The following generic PREMs have been sufficiently evaluated in German: HCAHPS, NORPEQ, PPE-15 and PEACS. For cancer care the EORTC IN-PATSAT32 and PSCC-G have been adequately assessed and can currently be recommended. We were unable to identify a surgery-specific PREM in the German language. However, even for the above mentioned generic and cancer care-specific PREMs certain deficits need to be considered before use. The HCAHPS for example, although showing sufficient psychometric properties in many areas, exhibits deficits in its cross-cultural validation into German (Table 7) [34]. Some of its demographic questions like “What is your race? Please select at least one.” were rated poorly by native German speakers [34] and refer to its development in a different sociocultural context. Therefore, an adaptation to the German-speaking sociocultural context seems necessary. The PPE-15, one of the most frequently used PREMs worldwide, exhibits poor structural validity, while covering many dimensions of PC (sufficient content validity). In addition, for many psychometric properties of the PPE-15 no data could be found. We cannot rule out that such data exist but has not been published by the Picker institutes or was not identified by our search. The NORPEQ is an extensively studied PREM with adequate psychometric properties. However, cross-cultural validation studies only exist for languages other than German, although it has been used in a non-validated German translations [43]. One of the most extensively evaluated generic PREMs is the PEACS questionnaire, that has been developed in German with involvement of patients. It is a comprehensive questionnaire with more than 50 questions covering many aspects of PC. Because of its length (Table 2) its intended use is as a reflection instrument for patients and as an assessment tool for providers rather than as benchmarking instrument. It is the only German generic PREM that covers not only in-hospital aspects of patient experience, but also the transition into out-patient care. Although the PEACS has sufficient psychometric properties in many areas, there is a lack of data for test-retest, inter-rater and intra-rater reliability. The PSCC-G is a cancer care specific PREM, that covers transition aspects of care. It has been built from a validated German translation of an English questionnaire with additional questions from other languages (Table 2). The PSCC-G scored adequately in many psychometric domains, but data on test-retest, inter-rater and intra-rater reliability are lacking.

### Limitations

The study has several limitations. First, the search was limited to generic, surgery-and cancer care specific PREMs, i.e., PREMs for other disciplines (e.g., internal medicine) as well as PREMs for specific diseases were excluded. These PREMs can be found in the excluded fulltext list (S7 Table). Another limitation could have been the search algorithm. The fact, that many German-language PREMs were identified by hand-searching rather than the database search, could be a hint that the search algorithm was not specific enough. However, the large number of identified and screened articles indicates that our search was broad. Furthermore, we were able to identify significantly more German-language PREMs than in previous reviews [8, 42]. Identifying PREMs in scientific databases is not easy. Contrary to PROMs no PREM-specific taxonomy (e.g., MeSH term) exists for PREMs in common medical databases. As pointed out, patient centredness and patient experience are only beginning to be clearly defined (S1 Fig and S2 Table). The delineation to other concepts like patient satisfaction is not always clear cut which makes building a search algorithm more difficult.

A main finding of the study is the lack of psychometric data for many of the included PREMs. Frequently we were unable to find appropriate studies in accessible databases. However, many PREMs have been developed and are implemented by independent or commercial institutions or healthcare agencies. These institutions are often not scientifically driven and might not publish all available psychometric data. Exception are the transparent development and publication of data by the U.S. *Agency for Healthcare Research and Quality (AHRQ)* (www.ahrq.gov/cahps/about-cahps/index.html) or the Swedish PREMs [18,44,45].

### Future research

If PC is supposed to be more than a declaration of intent of healthcare politicians, it will require the implementation of PREMs into everyday clinical practice via the following measures:

- As shown in our study, there is no comprehensive modular PREM system in the German language comparable to other countries [23]. Many areas of healthcare (e.g., surgery) are not covered with available German-language PREMs. Consequently, the development, translation and testing of new PREMs is necessary.
- The missing psychometric properties of currently available German-language PREMs need to be evaluated.
- Most of the PREMs currently available are paper-based versions (Table 2). For broad implementation and timely assessment in hospitals and doctoŕs offices electronic PREMs (ePREM) seem necessary. For this purpose, paper-based PREMs will need to be evaluated as digital versions and electronic systems will have to be developed and implemented that adhere to local data safety regulations. An integration into available hospital information systems is desirable, to facilitate the use in everyday clinical practice.
- It is unclear which conclusions should be drawn from the results of PREM (sub)scales. If providers adapt their service based on PREM results, there is little evidence-base to guide such changes [46]. Individualized local measures may be implemented, but there may also be standardized interventions, which can be tested in randomized-controlled trials which might improve aspects of PC and subsequently PREM results. More research is needed in this field.

There are two projects that should be mentioned in this context. First, the EORTC is currently developing and testing the PATSAT-33, a cancer-care specific PREM that will not only cover in-hospital patients, but also aspects of PC in out-patient settings as well as the transitional period [47]. A phase IV validation study in several European countries is underway including German-speaking countries. Second, the Hamburg-based ASPIRED project [48], is currently developing a German-language PREM, that will cover all aspects of PC according to Scholl et al. [5]. Both projects will close important evidence gaps.

### Conclusions

This systematic review and qualitative analysis of the psychometric properties of German-language PREMs, gives an overview of available generic and cancer-care specific PREMs. Results can be used to choose valid and reliable PREMs according to the above-mentioned recommendations. Furthermore, areas of future research are delineated.

## Supporting information

S2_PatientCentrednessDimesions

S3_PRISMAchecklist

S4_SearchAlgorithm

## Data Availability

All relevant data are within the manuscript and its Supporting Information files

## Author contributions

AMi and GS designed the study. EK performed the database searches in coordination with AMi. AMi and CDH screened titles and abstracts as well as full texts. AMi and CDH performed the data extraction. AMi and CDH performed the qualitative analyses. AMi and GS drafted the manuscript. All authors revised the manuscript critically. All authors read and approved the manuscript.

## Acknowledgements

The work is based on the Master thesis of AMi for the Master of Business Administration (MBA) in Healthcare Management at the Wirtschaftsuniversität Wien (Vienna University of Economics and Business) (2019-2021) for which GS acted as mentor and supervisor.

## Supporting information

**S1 Table.** Dimensions of patient centredness.

**S2 Table.** PRISMA Checklist

**S3 Text.** Search algorithm

**S4 Text.** Material and methods

**S5 Table.** Non-German PREMs

**S6 Table.** Content validity

