## Supplementary material for "Measuring patient centeredness with German language Patient-Reported Experience Measures (PREM) – a systematic review and qualitative analysis according to COSMIN": S2_PatientCentrednessDimesions

Supplement 2. Dimensions of patient centredness

|  | Scholl et al. (2014) | Wong et al. (2020) | Picker | Mead & Bower (2000) |
| --- | --- | --- | --- | --- |
| 1 | *patient as a unique person* | *patient preference* | *Respect for patient preferences* | *the `patient-as-person'* |
| 2 | *biopsychosocial persp*e*ctive* |  |  | *Biopsychosocial persp*e*ctive* |
| 3 | essential characteristics of the clinician |  |  | *the `doctor-as-person'* |
| 4 | patient involvement in care | *patient preference* | *Involvement in treatment decisions* | *sharing power & responsibility* |
| 5 | involvement of family and friends | *family & friends* | *involvement of family and friends* | *therapeutic alliance* |
| 6 | physical support | *physical support* | *Physical well-being, clean and safe environment* |  |
| 7 | emotional support | *emotional support* | Emotional support, empathy, respect |  |
| 8 | clinician-patient communication | *information & education* | Clear and understandable information und support for self-care | *sharing power & responsibility* |
| 9 | *patient empowerment* |  |  |  |
| 10 | patient Information |  |  | *therapeutic alliance* |
| 11 | access to care | a*ccess to care* | Fast access to reliable healthcare |  |
| 12 | integration of medical and non-medical care |  |  |  |
| 13 | coordination and continuity of care | *coordination of care* | continuity of care & well-arranged care transition |  |
|  |  | *continuity & transition of care* |  |  |
| 14 | teamwork and teambuilding |  |  | *therapeutic alliance* |
| 15 | clinician-patient relationship |  |  |  |
| 16 |  |  | Effective treatment by trustworthy healthcare personnel |  |

Different models emphasize different dimenions of patient centredness: Scholl et al. from Scholl et al. An integrative model of patient-centeredness - a systematic review and concept analysis. PloS One. 2014;9(9):e107828. Wong et al. from Wong, E., Mavondo, F. & Fisher, J. Patient feedback to improve quality of patient-centred care in public hospitals: a systematic review of the evidence. BMC Health Serv Res 20, (2020). Picker from BQS Institut: Picker Patient-centred care model. 2021. <https://www.bqs.de/picker-befragungen/51-patientenbefragung.php>. Mead & Bower from Patient-centredness: a conceptual framework and review of the empirical literature. Social Science & Medicine 51, 1087–1110 (2000).
