## Supplementary material for "Measuring patient centeredness with German language Patient-Reported Experience Measures (PREM) – a systematic review and qualitative analysis according to COSMIN": S4_SearchAlgorithm

**Supplement 4. Search algorithm**

The search algorithm for EBM Reviews - Cochrane Database of Systematic Reviews, EBM Reviews - Health Technology Assessment, Embase, Ovid MEDLINE(R) and APA PsycInfo was as follows:

(((((Patient* or Consumer*) adj (satisfaction or Experience* or Opinion* or Perspective*)).m_titl. or exp Patient Satisfaction/) and ((Questionnaire* or Instrument* or measure*).m_titl. or (Health care surveys/ or questionnaires/))) or exp Patient Reported Outcome Measures/) and (validity or reliability or repeatability).mp.

The search algorithm for CINAHL was as follows:

(((((Patient* or Consumer*) adj (satisfaction or Experience* or Opinion* or Perspective*)).m_titl. or exp Patient Satisfaction/) and ((Questionnaire* or Instrument* or measure*).m_titl. or (Health care surveys/ or questionnaires/))) or exp Patient Reported Outcome Measures/) and (validity or reliability or repeatability).mp.
