## Supplementary material for "Measuring patient centeredness with German language Patient-Reported Experience Measures (PREM) – a systematic review and qualitative analysis according to COSMIN": S5_Methods

**Supplement 5. Methods for the qualitative analysis of psychometric properties**

Methods are according to current COSMIN guidelines [1,2].

### 1.1 Assessment of content validity

Content validity describes in how far the content of a PREM is an adequate reflection of the underlying construct. In this case the construct is patient-centredness. The domains of content validity have been described in Table 3 and S2 Table. Rating of content validity was carried out as recommended by the COSMIN study group [1]. For each German-language PREM the following questions were answered:

|  | Relevance |
| --- | --- |
| 1 | Are the included items relevant for the construct of interest? |
| 2 | Are the included items relevant for the target population of interest? |
| 3 | Are the included items relevant for the context of use of interest? |
| 4 | Are the response options appropriate? |
| 5 | Is the recall period appropriate? |
|  | Comprehensiveness |
| 6 | Are no key concepts missing? |
|  | Comprehensibility |
| 7 | Are the PROM instructions understood by the population of interest as intended? |
| 8 | Are the PROM items and response options understood by the population of interest as intended? |
| 9 | Are the PROM items appropriately worded? |
| 10 | Do the response options match the question? |

In addition the quality of methods fort he development of the PREM was rated. Again the criteria of the COSMIN group were used [1].

### 1.2 Analysis of structural validity

In the next steps the internal strucutre of the PREM was analysed. To this end the following 3 psychometric properties were evaluated: a.) structural validity, b.) internal consistency and c.) measurement invariance / cross-cultural validity [1]. Structural validity was defined as the degree to with a PREM results is an adequate reflection of the underlying concept [3]. Each German PREM was evaluated whether it was a reflective or formative questionnaire because analysis of structural validity is meaningful only for reflective questionnaires [4]. In the next step, it was analysed whether the study analyses the unidimensionality of a PREM or other aspects of structural validity. If the scale or subscale of PREM is unidimensional, all questions measure the same concept and will therefore change in the same direction if the underlying concept changes. Structural validity of a specific PREM will be assessed with the following questions from the COSMIN handbook [4]:

1.) Is the PREM based on a reflective model? (yes/no)
2.) Does the study investigate the unidimensionality of a construct? (yes/no)
3.) Statistical methods:
    a. In case of *Classical Test Theory*: was an explorative or confirmatory factor analysis performed?
       i. very good: Confirmatory factor analysis performed
       ii. : adequate: explorative factor analysis performed.
       iii. Inadequate: neither confirmatory nor exploratory factor analysis performed.
       iv. Not applicable.
    b. For IRT/Rasch: does the chosen model fit to the research question?
       i. very good: chosen modelfits well to the researchquestion
       ii. adequate: Assumable that the chosen model fits well to the research question
       iii. Doubtful: Doubtful if the chosen model fits well to the research question
       iv. inadequate: Chosen model does not fit to the research question
       v. Not applicable
4.) Was the sample size included in the analysis adequate (see Table 1 main manuscript)
5.) Were there any other important flaws in the design or statistical methods of the study?
    i. very good: No other important methodological flaws
    ii. doubtful: Other minor methodological flaws (e.g. rotation method not described)
    iii. inadequate: Other important methodological flaws
6.) Overall rating of structural validity (see Table 1 main manuscript)

### 1.3 Analysis of internal consistency

„Internal consistency refers to the degree of interrelatedness among the items and is

often assessed by Cronbach’s alpha …. For an appropriate interpretation of the internal consistency parameter, the items together should form a unidimensional scale or subscale” [4].

Internal consistency was evaluated with following questions from the COSMIN handbook [4]:

1.) Does the scale consist of effect indicators, i.e. is it based on a reflective model (yes/no)
2.) Was an internal consistency statistic calculated for each unidimensional scale or subscale separately?
    i. Very good: Internal consistency statistic calculated for each unidimensional scale or subscale.
    ii. Doubtful: unclear whether scale or subscale is unidimensional
    iii. Inadequate: Internal consistency statistic NOT calculated on unidimensional scale
3.) Statistical methods:
    a. For continuous scores: Was Cronbach’s alpha or omega calculated?
       i. Very good: Cronbach`s Alpha or Omega calculated.
       ii. doubtful: only item-total correlations calculated.
       iii. inadequate: no Cronbach`s Alpha and no item-total correlations calculated
       iv. not applicable
    b. for dichotomous scores: Was Cronbach`s Alpha or KR-20 (Kuder-Richardson Formula 20) calculated?
       i. Very good: Cronbach`s Alpha or KR-20 calculated
       ii. Doubtful: only item-total correlations calculated
       iii. inadequate: no Cronbach`s Alpha or KR-20 and no item-total correlations calculated
       iv. not applicable
4.) Overall rating of structural validity (see Table 1 main manuscript)

### 1.4 Analysis of measuremtn invariance / Cross-cultural validity

*Measurement invariance / cross-cultural validity* describes to what degree the measurements of a PREM (total scale or subscales) differ, when it is translated or used in another clinical or cultural context. *Cross-cultural validity* is interpreted broadly by current COSMIN guidelines, and includes using the PREM in another age group or patient cohort [4]. For this reason we use the term *measurement invariance* rather than *cross-cultural validity* [4]. Measurement invariance is frequently analysed using *Measurement Invariance* (MI) or the *differential item functioning* (DIF) [2]. Analysis of the measurement invariance is important for internaional PREMs that have been translated into German. In our study measurement invariance was evaluated as outlined in the COSMIN guidelines [4].

### 1.5 Analysis of reliability

“Reliability refers to the proportion of the total variance in the measurements which is

due to ‘true’ differences between patients” [4]. The ’true’ differences would be the average score that would be obtained if the scale was administered an infinite number of times to the same person. „It refers only to the consistency of the score, and not to its accuracy“ [5]. “An important assumption made in a reliability study (…) is that

patients are stable on the construct to be measured between the repeated measurements “ [4]. In our study reliability was evaluated as outlined in the COSMIN guidelines [4].

### 1.6 Analysis of measurement error

„Measurement error refers to the systematic and random error of an individual patient’s

score that is not attributed to true changes in the construct to be measured” [4]. Thus, it can be viewed as the opposite of reliability. In our study measurement error was evaluated as outlined in the COSMIN guidelines [4].

### 1.7 Analysis of criterion validity

„Criterion validity refers to the degree to which the scores of a PROM are an adequate

reflection of a ‘gold standard’“ [4]. The current gold standard needs to be determend for each PREM scale or subscale and depends on the content domain of the PREM (Table 3 main manuscript). As outlined in the main manuscript no gold standard for patient centredness has yet been defined. Therefore, analysis of criterion validity is only possible for certain subscales. If the PREM and the gold standard are continuous scores, the calculation of correlations is the adequate method [4]. If the PREM is a continuous score, but the gold standard is dichotomous score, the calculation of the *Area Under the receiver operating Curve* (AUC) is the most adequate statistical method to calculate the criterion validity. If both scales are dichotomous, calculation of sensitivity and specificity are the most appropriate method [4]. In our study criterion validity was evaluated as outlined in the COSMIN guidelines [4].

### 1.8 Analysis of hypothesis testing for construct validity

„Hypotheses testing for construct validity refers to the degree to which the scores of a PROM are consistent with hypotheses (…)based on the assumption that the PROM validly measures the construct to be measured” [4]. “Hypotheses testing is an ongoing, iterative process” [4]. Many different hypotheses can be tested. The more hypotheses are tested and the more specific the hypotheses are, the more evidence can be generated of construct validity. “In general, these hypotheses concern comparisons with other outcome measurement instruments (*convergent validity*), or differences in scores between ‘known’ groups (*known-groups validity*)” [4]. In our study hypothesis testing for construct validity was evaluated as outlined in the COSMIN guidelines [4].
