## Supplementary material for "Measuring patient centeredness with German language Patient-Reported Experience Measures (PREM) – a systematic review and qualitative analysis according to COSMIN": S6_Table

**Supplement 6 Table**. Study characteristics of all non-German Patient-Reported Experience Measures (PREMs) identified in the search. A) generic; B.) surgery-specific and C.) cancer care-specific PREMs.

A.) **Generic PREMs**

|  | Author | Ref. | Year | Acronym | Country | Description | Method (timepoint) of assessment | Number of items | Domains | Comments |
| --- | --- | --- | --- | --- | --- | --- | --- | --- | --- | --- |
| 1 | Fernstrom KM, Shippee ND, Jones AL, Britt HR | BMC Palliat Care. 2016;15(1):99 | 2016 | N/A | USA | “For patients with serious chronic illness“, „Aims to address shortcomings of HCAPHS, which ignores the broader context of care delivery via teams, care encounters across settings, health declines, life transitions“ | paper-based (not specified) | 21 | 1. Care Team 2. Communication 3. Care Goals |  |
| 2 | Tian CJ, Tian Y, Zhang L. | J Huazhong Univ Sci Technolog Med Sci. 2014;34(2):9. | 2014 | PEES-50 | China | To assess medical service quality in the Chinese context | paper-based (at discharge) | 50 | 1. tangibility 2. reliability 3. responsiveness 4. assurance 5. empathy 6. continuity |  |
| 3 | Bruyneel L, Tambuyzer E, Coeckelberghs E, et al. | Int J Environ Res Public Health. 2017;14(11):14. | 2017 | N/A | Belgium | “Standardized questionnaire for public reporting in Flanders”; in Flemish | paper-based and electronic  (“at the time of discharge, but hospitals may also opt for administration after discharge”) | 28 | 1. Preparing for hospital stay 2. Information and communication 3. Coordination 4. Respect 5. Privacy 6. Safe Care 7. Pain management 8. Participation |  |
| 4 | González N, Quintana JM, Bilbao A, et al. | Int J Qual Health Care. 2005;17(6):465‐472. | 2005 | N/A | Spain | “Inpatient satisfaction questionnaire to evaluate the health care received by patients admitted to several hospitals”. “Evaluation of possible predictors of patient satisfaction in relation to socio-demographic variables, history of admission, and survey logistics” | paper-based (2 weeks after discharge); 2 reminders within 2 weeks | 34 | 1. information and 2. communication with doctors (12 items) 3. nursing care (8 items) 4. comfort (6 items) 5. visiting (4 items) 6. privacy (2 items) 7. cleanliness (2 items) |  |
| 5 | Labarère J, Fourny M, Jean‐Phillippe V, Marin‐Pache S, Patrice F. | Int J Health Care Qual Assur. 2004;17(1):17‐25. | 2004 | N/A | France | “Assessing patient experience in French hospitals…”. “But a validated French-language questionnaire is currently lacking and translating foreign scales seems inappropriate because of substantial structural differences between health care systems” | paper-based (2-4 weeks after discharge) | 29 | 1. Medical information 2. Nursing Care 3. Living arrangements 4. Discharge Management 5. Coordination 6. Physician care 7. Convenience |  |
| 6 | Labarere J, Francois P, Auquier P, Robert C, Fourny M. | Int J Qual Health. 2001; 13(2) | 2001 | N/A | France | „to measure inpatient satisfaction“ | paper-based (2-4 weeks after discharge) | 30 | 1. Nursing care 2. Communication 3. Discharge planning/continuity 4. Physician care 5. Living arrangements 6. Convenience | Tested in 30 patients |
| 7 | Pettersen KI, Veenstra M, Guldvog B, Kolstad A. | J Qual Health Care. 2004;16(6):11 | 2004 | PEQ | Norway | “…for measuring patient experiences of hospital care, applicable both in quality improvement locally and for national surveillance” | paper-based (6 weeks after discharge; reminder after 4 weeks) | 35 | 1. Information on future complaints 2. Nursing services, 3. Communication, 4. Information examinations 5. Contact with next-of-kin, 6. Doctor services 7. Hospital and equipment, 8. Information medication, 9. Organization 10. General satisfaction |  |
| 8 | Sjetne IS, Bjertnaes OA, Olsen RV, Iversen HH, Bukholm G. | BMC Health Serv Res. 2011;11:11. | 2011 | GS‐PEQ  (generic short Patient Experience Question-naire) | Norway | “Selection of ten generic core questions that cover the essential dimensions of users’ experiences with the services provided across a range of specialist health care service” | paper-based (after discharge, 2 reminder) | 10 | 1. Outcome, 2. clinician services, 3. user involvement, 4. incorrect treatment, 5. information, 6. organisation, 7. accessibility | Short version of the PEQ (Pettersen et al.) |
| 9 | Sullivan PJ, Harris ML, Doyle C, Bell D | BMJ Qual Saf. 2013;22(8):690‐696. | 2013 | NHS adult-inpatient survey | England | To determine whether validity of AIP (Adult In-Patient Survey) is retained at a suborganisational level | paper-based (after discharge) | 30 | 1. Nurses 2. Doctors 3. Hospital and ward 4. Care and treatment 5. Operations and procedure 6. Leaving hospital 7. Overall | Further references in:  1.) Boyd J. The 2006 inpatients importance study. Oxford, Picker Institute Europe: The Acute Co-ordination Centre for the NHS Acute Patient Survey Programme; 2007.  2.) DeCourcy A, West E, Barron D. The National Adult Inpatient Survey conducted in the English National Health Service from 2002 to 2009: how have the data been used and what do we know as a result? BMC Health Serv Res. 2012;12:71  3.) Picker Institute Europe. Guidance manual for the NHS Adult Inpatient Survey 2012. Oxford: Picker Institute Europe; 2012;  4.) Sizmur S, Redding D. Core domains for measuring inpatients’ experience of care. Oxford: Picker Institute Europe; 2012. |
| 10 | Wong EL, Coulter A, Cheung AW, Yam CH, Yeoh EK, Griffiths S. | BMC Health Serv Res. 2013;13. | 2013 | HKIEQ  (Hong Kong Inpatient Experience Questionnaire) | Hong Kong | to develop a tool for assessing patient experience with inpatient care in public hospitals in Hong Kong | paper-based (48 hours to 1 month after discharge) | 58 | 1. Information provision 2. Care & involvement in decision making 3. Physical & emotional needs 4. Coordination of care 5. Respect & privacy 6. Environment & facilities 7. Handling patient feedback 8. Overall care of healthcare professionals & Quality of Care |  |
| 11 | Wong ELY, Coulter A, Hewitson P, et al. | PLoS ONE. 2014;10(4):12. | 2014 | SF-HKIEQ | Hong Kong | “To develop a short-form version of the HKIEQ.” | paper-based (48 hours to 1 month after discharge) | 18 | 1. Hospital Staff 2. Patient Care & Treatment 3. Information on Leaving Hospital 4. Overall Impression | Short-form of the HKIEQ |
| 12 | Bamm EL, Rosenbaum P, Stratford P. | Int J Qual Health Care. 2010;22(4):302‐309. | 2010 | MPOC-A  (measure of processes of care for adults) | Canada | Validation “To assess the psychometric properties of the measure of processes of care for adults (MPOC-A)” | paper-based (“6 months after planned joint replacement surgery”) | 34 | 1. Enabling and Partnership 2. Providing General Information 3. Providing Specific Information 4. Coordinated and Comprehensive Care 5. Respectful and Supportive Care | Administered along with 8-items CSQ (client satisfaction questionnaire) |
| 13 | Benson T, Potts HWW. | BMC Health Serv Res. 2014;14(1):499 | 2014 | HowRwe | England | Development and validation of the howRwe questionnaire | paper-based or tablet  (timepoint not specified) | 4 | 1. Kindness 2. Organisation 3. Communication 4. Punctuality | Further references:  Hendriks SH, Rutgers J, van Dijk PR, et al. Validation of the howRu and howRwe questionnaires at the individual patient level. BMC Health Serv Res. 2015;15:8. |
| 14 | Malott DL, Fulton BR, Rigamonti D, Myers S. | J Surv Stat Methodol. 2017;5(3):11. | 2017 | N/A | Saudi Arabia, United Arab Emirates | Generic in-hospital PREM | paper-based  (timepoint not specified) | 34 | 1. Admission 2. Food & accommodation 3. Nurses 4. Personal aspects of care 5. Physician 6. Overall assessment |  |
| 15 | Webster TR, Mantopoulos J, Jackson E, et al. | Int J Qual Health Care. 2011;23(3):11 | 2011 | I-PAHC | Ethiopia | Low-income setting. Completion after first day of admission. | paper-based  (First day aafter admission) | 25 | 1. Nurse 2. Communication 3. Doctor communication 4. Physical environment 5. Pain management & medication 6. Symptom communication |  |
| 16 | Boge RM, Haugen AS, Nilsen RM, Harthug S. | PLoS One. 2018 Nov 7;13(11):e0206904. | 2018 | DICARES | Norway | „develop and validate a survey instrument feasible for measuring quality related to the discharge process based on elderly patients’ experiences“ | paper-based (30 days after discharge) | 10 | 1. Coping after discharge 2. Participation in discharge planning 3. Adherence to treatment |  |
| 17 | Casu G, Gremigni P, Sommaruga M. | Patient Educ Couns. 2019 Jan;102(1):126-133. | 2019 | PPIQ | Italy | to confirm the psychometric properties of the PPRQ in its patient form, named  Professional-Patient Interaction Questionnaire (PPIQ)  to confirm the psychometric properties of the PPRQ in its patient form, named  Professional-Patient Interaction Questionnaire (PPIQ)  to confirm the psychometric properties of the PPRQ in its patient form, named  Professional-Patient Interaction Questionnaire (PPIQ)  “…to assess patient centered care from the patient's perspective” | on-site, a paper-and-pencil questionnaire about the most recent  Encounter with a professiona  paper-based (before discharge) | 16 | 1. Effective communication 2. Interest in the patient's agenda 3. Empathy 4. Patient involvement in care | for in- and out-patients |
| 18 | Christalle E, Zeh S, Hahlweg P, Kriston L, Härter M, Scholl I. | BMJ Open. 2018 Oct 21;8(10):e025896. | 2018 | ASPIRED | Germany | “to assess relevance of dimensions of patient centredness from the patients’ perspective”  “to develop and psychometrically test a core set of patient-reported experience measures (PREMs) assessing patient centredness” |  |  |  | Study protocol. German PREM under development. |
| 19 | Eubank BH, Lafave MR, Mohtadi NG, Sheps DM, Wiley JP. | Prim Health Care Res Dev. 2019 Jun 11;20:e47. | 2019 | HAPSQ | Canada | “Validation of a tool to assess patient  satisfaction, waiting times, healthcare  utilization, and cost” | Web-based questionnaire | 29 | 1. Accessibility 2. Acceptability 3. Efficiency 4. Appropriateness | Measures discharge process from hospital. |
| 20 | Graumlich JF, Novotny NL, Aldag JC. | J Hosp Med. 2008 Nov-Dec;3(6):446-54. | 2008 | B-PREPARED | USA | “… to measure preparedness for hospital discharge from the patient’s perspective” | telephone (1 week after discharge) | 11 | 1. Self-care Information for Medications and Activity 2. Equipment and Services 3. Confidence | Measures readiness of discharge |
| 21 | Zhang J, Zhou F, Ge X, Ran X, Li Y, Chen S, Dai X, Chen D, Jiang B. | Patient Prefer Adherence. 2018 Nov 29;12:2527-2536. | 2018 | IPSQ | China | To verify the reliability and validity of the indicator system for evaluating satisfaction of outpatients and  inpatients in Chinese hospitals | „mobil terminal system“ (before discharge) | 50 | 1. doctor–patient relationship 2. doctor–patient communication 3. medical service 4. auxiliary service 5. environmental sanitation 6. medical ethics 7. procedure signs | Validation in Chinese |
| 22 | Larsson G, Larsson BW, Munck IME. | Scand J Caring Sci. 1998;12:111–8.; | 1998 | QPP  (Quality from the Patients'  Perspective) | Sweden | Generischer PREM für stationäre Patienten für das schwedische Gesundheitssystem | paper-based (at discharge) | 68 | 1. Medical-technical competence 2. Physical technical conditions 3. Personal necessities Characteristics 4. Identity-orientated approach 5. Situation 6. Participation 7. Commitment 8. Socio-cultural atmosphere 9. Positive treatment of significant others | Further references:  1.) Wilde B, Larsson G, Larsson M, Starrin B. Quality of care: development of a patient-centred questionnaire based on a grounded theory model. Scand J Caring Sci. 1994;8:39–48.;  2.) Wilde B, Starrin B, Larsson G, Larsson M. Quality of care from a patient perspective: a grounded theory study. Scand J Caring Sci. 1993;7:113–20 |
| 23 | Scottish Government | Scottish Inpatient Patient Experience Survey 2010, Volume 2: Technical Report. (http://www.scotland.gov.uk/Publications/2010/  09/30111425/0). | 2010 | SIPES  (Scottish Inpatient Patient Experience Survey) | Scotland | “(1) Gain an understanding of the experiences of adult patients receiving services at NHS hospitals in Scotland”  “(2) Provide NHS Boards and NHS sites with information about areas of best practice and areas for improvement”  “(3) Determine the key drivers for positive inpatient experience within Scotland;”  “(4) Explore if and how differences exist in terms of experiences between patients of different (age) groups, gender…” | paper-based (before discharge) | 38 | 1. Admission to hospital 2. The hospital and the ward 3. Care and treatment 4. Staff 5. Leaving hospital 6. Overall experience | Further references:  Scottish Government. Scottish Inpatient Patient Experience Survey 2012, Volume 2: Technical Report. (http://www.scotland.gov.uk/Publications/2012/08/5616/0). |
| 24 | Rao KD, Peters DH, Bandeen-Roche K. | Int J Qual Health C. 2006;18:414–21 | 2006 | PPQ  (Patient Perceptions of Quality) | India | ”To develop a reliable and valid scale to measure in-patient and outpatient perceptions of quality in India” and  “To identify aspects of perceived quality which have large effects on patient satisfaction.” | Interview (in-hospital and out-patients) | 16 | 1. medicine availability 2. medical information 3. staff behavior 4. doctor behavior 5. hospital infrastructure | Visual representation of the Likert scale |
| 25 | Wei J, Wang XL, Yang HB, Yang TB. | PLoS One. 2015 Dec 11;10(12):e0144785. | 2015 | N/A | China | “To develop a reliable and practical questionnaire for the assessment of inpatients’ satisfaction in Chinese people” | paper-based (during in-hospital stay) | 28 | 1. Doctors’ care quality 2. Nurses’ care quality 3. Quality of the environment 4. facilities’ 5. Global quality |  |
| 26 | Wang X, Chen J, Yang Y, Burström B, Burström K. | Int J Equity Health. 2021 Jan 7;20(1):25. | 2021 | PREM-CCH  (patient-reported experience measure for care in Chinese hospitals) | China | Development in China | Interview | 22 (out-patient)  11 (in-hospital) | Out-patients:   1. Communication and Information 2. Professional competence 3. Medical costs 4. Efficiency 5. Hospital recommendation   in-hospital patients:   1. Communication and information 2. Professional competence 3. Medical costs 4. Efficiency 5. Health outcomes 6. Hospital recommendation |  |

B.) **Surgery-specific PREMs**

|  | Author | Ref. | Year | Acronym | Country | | Description | Method (timepoint) of assessment | Number of items | | Domains | Comments |
| --- | --- | --- | --- | --- | --- | --- | --- | --- | --- | --- | --- | --- |
| 1 | Cheung CS, Bower WF, Kwok SC, van Hasselt CA. | Contributors to Surgical In-patient Satisfaction—Development and Reliability of a TargetedInstrumentAsian J Surg. 2009;32(3):143‐150. | 2009 | HK2Happ  (Hong Kong Index of Inpatient Happiness) | Hong Kong | The HK2Happ was developed in Hong Kong for adult surgical patients. | | paper-based  (timepoint not defined) | 40 | 1.) Patient-clinician communication 2.) Patient information 3.) Physical support 4.) Emotional support 5.) Essential characteristics of the clinician 6.) Patient-Clinician relationship 7.) Teamwork | | Further references:  Wendy Fiona Bower, Catherine S.K. Cheung, Eric M.C. Wong, Ping-Yin Lee, Charles Andrew Van Hasselt, Surgical patient satisfaction in Hong Kong: Validation of a new instrument, Surgical Practice, 21 October 2009 |
| 2 | Schmocker RK, Cherney Stafford LM, Siy AB, Leverson GE, Winslow ER. | Understanding the determinants of patient satisfaction with surgical care using the Consumer Assessment of Healthcare Providers and Systems surgical care survey (S-CAHPS). Surgery. 2015 Dec;158(6):1724-33. | 2015 | S-CAHPS  (Consumer Assessment of Healthcare Providers and Systems surgical care survey) | USA | The Surgical CAHPS was developed for adulat surgical patients. It can be used for in- and out-patients. | | paper-based  (timepoint not specified) | 47 | 1.) Patient-clinician communication 2.) Patient information 3.) Empowerment 4.) Patient involvement 5.) Patient-Clinician relationship 6.) Patient as a unique person 7.) Physical support 8.) Coordination and transition of care 9.) Teamwork | | www.ahrq.gov/cahps/surveys-guidance/surgical/index.html |

C.) **Cancer care-specific PREMs**

|  | Author | Ref. | Year | Acronym | Country | Description | Method (timepoint) of assessment | Number of items | Domains | Comments |
| --- | --- | --- | --- | --- | --- | --- | --- | --- | --- | --- |
| 1 | Iversen HH, Holmboe O, Bjertnæs ØA. | BMJ Open. 2012;2(5):15 | 2012 | CPEQ  (Cancer Patient Experiences Questionnaire) | Norway | For in-hospital cancer patients independent of treatment discipline. | paper-based  (timepoint not specified) | 37 | 6 subscales for outpatients::   1. Nurse contact 2. Doctor contact 3. Information 4. Organisation 5. Patient safety 6. Contact with next of kin   One addition subscale for in-hospital patients:  7. Hospital standard | National assessment of *patient experience* in Norway |
| 2 | Abel GA, Gomez-Cano M, Pham TM, Lyratzopoulos G. | Reliability of hospital scores for the Cancer Patient Experience Survey: analysis of publicly reported patient survey data. BMJ Open. 2019 Jul 24;9(7):e029037 | 2019 | CPES  (Picker Cancer Patient Experience Survey) | Great Britain | “To assess the degree to which variations in publicly reported hospital scores arising from the English Cancer Patient Experience Survey (CPES) are subject to  Chance“  Reliability measurement of the CPES for cancer patients | paper-based or online (after discharge or after out-patient care) | 61  (v. 2019) | 1. Coordination of care 2. Access to care 3. Patient information 4. Clinician-patient communication 5. Involvement of family & friends 6. Emotional support 7. Physical support 8. Involvement in care 9. Patient empowerment 10. Teamwork 11. Patient as a unique person 12. Essential characteristics of the clinician | Picker questionnaire. More information:  1.) Quality Health. Guidance material and survey material. 2016. http:// www.ncpes.co.uk/reports/2016-reports/guidance-material-and- survey-materials/3211-2016-cancer-survey-guidance/file  2.) [www.oecd.org/els/health-systems/39493930.pdf](http://www.oecd.org/els/health-systems/39493930.pdf)  3.) Pham TM, Abel GA, Gomez-Cano M, Lyratzopoulos G. J Med Internet Res. 2019 May 2;21(5):e11855. |
| 3 | Brédart A, Beaudeau A, Young T, Moura De Alberquerque Melo H, Arraras JI, Friend L, Schmidt H, Tomaszewski KA, et al. ; EORTC Quality of Life Group. | The European organization for research and treatment of cancer – satisfaction with cancer care questionnaire: revision and extended application development. Psycho-Oncology 26(3): 400-404. | 2017 | EORTC PATSAT-C33 | Europe | The questionnaire contains items about the coordination and transition of care | paper-based (not specified) | 33 | Provisional list of issues by domain:  Doctors:   1. Technical skills 2. Information 3. Interpersonal skills 4. Availability   Nurses/radiotherapy technicians:   5. Technical skills 6. Information 7. Interpersonal skills 8. Availability   Services and care organization:   9. Continuity 10. Coordination 11. Interpersonal skills 12. Information provision 13. Waiting time 14. Privacy 15. Access   Outpatient module:   16. Continuity 17. Convenience 18. Waiting time 19. Access 20. Transition | This questionnaire is being evaluated in a multinantional phase IV study. A German translation is in preparation.. |
| 4 | Bredart A, Razavi D, Robertson C, Didier F, Scaffidi E, de Haes JC. | A comprehensive assessment of satisfaction with care: preliminary psychometric analysis in an oncology institute in Italy. Ann Oncol 1999;10:839–846. | 1999 | CASC  (Comprehensive Assessment of Satisfaction with Care) | Italy | Validation of the Italian translation of the CASC in oncological in- and out-patients | paper-based (after discharge or after out-patient care) | 50 | 1. Doctors 2. Nurses 3. Access/comfort 4. Care organization 5. General satisfaction | Further references:   1.) Bredart A, Razavi D, Robertson C, et al. A comprehensive assessment of satisfaction with care: preliminary psychometric analysis in French, Polish, Swedish, and Italian oncology patients. Patient Educ Couns 2001;43:243–252 2.) Kritsotakis, G., Koutis, A. D., Kotsori, A., Alexopoulos, C. G., & Philalithis, A. E. (2009). Measuring patient satisfaction in oncology units: interview-based psychometric validation of the ‘Comprehensive Assessment of Satisfaction with Care’ in Greece. European Journal of CancerCare, 19 (1), 45–52 |
| 5 | Booij JC, Zegers M, Evers PM, Hendriks M, Delnoij DM, Rademakers JJ. | Improving cancer patient care: development of a generic cancer consumer quality index questionnaire for cancer patients. BMC Cancer 2013;13:203. | 2013 | CQI‐CC (Consumer Quality Index Cancer Care) | The Nether-lands | Development of a generic questionnaire for oncological patients | Online  (Independent of timepoint of treatment. For in- and out-patients) | 99 | 12 Subscales:   1. Personal attention during aftercare 2. Cooperation & communication between healthcare professionals 3. Freedom of choice 4. Skills and cooperation of healthcare professionals 5. Psychosocial guidance 6. Other investigations and treatments 7. Information during treatment 8. Continuity of care by healthcare professional/side effects and complaints. 9. Patient-centered approach by doctors 10. Patient-centered approach by nurses 11. Information at completion of treatment 12. Transfer to other healthcare professionals |  |
| 6 | Peipert JD, Beaumont JL, Bode R, Cella D, Garcia SF, Hahn EA. | Development and validation of the functional assessment of chronic illness therapy treatment satisfaction (FACIT TS) measures. Qual Life Res 2014;23(3):815–824. | 2014 | FACIT‐TS (Functional Assessment of Chronic Illness Therapy Treatment Satisfaction) | USA | Development and validation of a FACIT questionnaire for chronic illnesses. | Multi-stage mixed method: Interview and paper-based | 26 | FACIT-TS (PS):   1. Physician Communication 2. Treatment Staff Communication 3. Technical Competence 4. Confidence and Trust 5. Nurse Communication | Consists of 2 parts:  (1) the FACIT TS-general (G)  (2) the FACIT TS-patient satisfaction (PS) |
| 7 | Cheater FM, Preston C, Wynn A, Hearnshaw H, Baker R. | Patients' views of cancer services: development of a questionnaire for accreditation. Eur J Oncol Nurs 1999;3(2):72–82 | 1999 | PVCS  (Patient Views of Cancer Services) | Great Britain | Questionnaire fort he accreditation of cancer centres. | paper-based (recall-period 6 months) | 77 | 1. Information and support 2. Interpersonal skills of health professionals 3. Speed of referrals 4. Environment of care 5. Carer´s perpective | Recall-Bias |
| 8 | Malin, J. L., Ko, C., Ayanian, J. Z., Harrington, D., Nerenz, D. R., Kahn, K. L., et al. | Understanding cancer patients' experience and outcomes: development and pilot study of the Cancer Care Outcomes Research and Surveillance patient survey. Support Care Cancer. Aug;14(8):837-48. | 2006 | CCORS  (Cancer Care Outcomes Research and Surveillance patient survey) | USA | Questionnaire to assess the experiences and outcomes of patients with colorectal or lung cancer | computer-assisted telephone interviewing (within 4 months after diagnosis) | unclear | 1. Organisation of patient care 2. Access to and navigation through the healthcare system 3. Allocation of a “key contact” person 4. Recognition and understanding of medical team roles 5. Effective communication and cooperation amongst the multidisciplinary team and other health service providers 6. Delivery of services in a   complementary and timely manner   7. Needs assessment 8. Sufficient and timely information for the patient |  |
| 9 | Young, J. M., Walsh, J., Butow, P. N., Solomon, M. J., & Shaw, J. | Measuring cancer care coordination: development and validation of a survey for patients. BMC Cancer, 11, 298 | 2011 | CCCQ  (cancer care coordination questionnaire) | Australia | Questionnaire to assess cancer care coordination.  Analyses of psychometric properites in two patient cohorts | paper-based (not specified ) | 20 | 8 subscales:   1. Organisation of patient care 2. Access to and navigation through the healthcare system 3. Allocation of a “key contact” person 4. Recognition and understanding of medical team roles 5. Effective communication and cooperation amongst the multidisciplinary team and other health service providers 6. Delivery of services in a complementary and timely manner 7. Needs assessment 8. Sufficient and timely information for the patient | Translated and validated in Arabic and Chinese |
| 10 | Wind A, Roeling MP, Heerink J, Sixma H, Presti P, Lombardo C, van Harten W. P | Piloting a generic cancer consumer quality index in six European countries. BMC Cancer. Sep 2;16(1):711. | 2016 | ECCQI  (European Cancer Consumer Quality Index) | Europe | International validation of the Dutch original questionnaire | Paper-based and online (timepoint not specified) | 45 | 10 categories:   1. Accessibility 2. Organization 3. Hospitalization 4. Safety 5. Attitude of HP 6. Communication and information 7. Own input 8. Coordination 9. Supervision and support 10. Rounding off the treatment | No translation and validation in German  Further references in:  Wind A, Hartman ED, Van Eekeren RRJP, Wijn RPWF, Halámková J, Mattson J, Siesling S, van Harten WH. Validating a generic cancer consumer quality index in eight European countries, patient reported experiences and the influence of cultural differences. BMC Cancer. 2021 Mar 6;21(1):231. |
| 11 | Evensen CT, Yost KJ, Keller S, Arora NK, Frentzel E, Cowans T, Garfinkel SA. | Development and Testing of the CAHPS Cancer Care Survey. J Oncol Pract. 2019 Nov;15(11):e969-e978. | 2019 | CAHPS  Cancer Care Survey | USA | Systematic assessment of the experience of cancer patients | paper-based and Web-Mail (after treatment -surgery, radiation or chemotherapy) | 38 | Two global ratings:   1. Cancer center overall 2. Therapy-specific team   8 composite measures:   1. Supporting patient self-management 2. keeping patients informed 3. providing care and information when needed 4. shared decision-making 5. access to care 6. communication between providers and patients 7. care coordination 8. courteous office staff | CAHPS Cancer Care Survey. https://www.ahrq.gov/cahps/surveys-guidance/cancer/index.html |
