## Supplementary material for "Measuring patient centeredness with German language Patient-Reported Experience Measures (PREM) – a systematic review and qualitative analysis according to COSMIN": S7_Table

**Supplement 8.** Excluded full text articles.

|  | Autor | Ref. | Kürzel | Ausschlussgrund | Kommentar |
| --- | --- | --- | --- | --- | --- |
| 1 | Sitzia J, Wood N. | Development and evaluation of a questionnaire to assess patient satisfaction with chemotherapy nursing care. Eur J Oncol Nurs 1999;3:126–140 | N/A | No multidimensional PREM | Only covers chemotherapy associated nusing-care |
| 2 | Panouilleres M, Anota A, Nguyen TV, et al. | Evaluation properties of the French version of the OUT‐PATSAT35 satisfaction with care questionnaire according to classical and item response theory analyses. Qual Life Res 2014 | OUT-PATSAT35 | PREM for outpatient sector | Outpatients only |
| 3 | Arraras JI, Illarramendi JJ, Viudez A, et a | The cancer outpatient satisfaction with care questionnaire for chemotherapy, OUT‐PATSAT35 CT: a validation study for Spanish patients. Support Care Cancer 2012;20:3269–3278. | OUT‐PATSAT35-CT | PREM for outpatient sector | Outpatients only |
| 4 | Arraras JI, Rico M, Vila M, et al. | The EORTC cancer outpatient satisfaction with care questionnaire in ambulatory radiotherapy: EORTC OUT‐PATSAT35 RT. Validation study for Spanish patients. Psycho‐Oncology 2010;19(6):657–664 | OUT‐PATSAT35-RT | PREM for outpatient sector | Outpatients only |
| 5 | Poinsot R, Altmeyer A, Conroy T, et al. | Multisite validation study of questionnaire assessing out‐patient satisfaction with care questionnaire in ambulatory chemotherapy or radiotherapy treatment. Bull Cancer 2006;93:315–327 | N/A | PREM for outpatient sector | Outpatients only |
| 6 | Sitzia J, Wood N. | Development and evaluation of a questionnaire to assess patient satisfaction with chemotherapy nursing care. Eur J Oncol Nurs 1999;3:126–140 | WCSQ | No generic, surgery- or cancer care-specific PREM | PREM only for adverse events of chemotherapy |
| 7 | Boyes, A., Girgis, A., & Lecathelinais, C. | Brief assessment of adult cancer patients’ perceived needs: development and validation of the 34-item Supportive Care Needs Surve (SCNS-SF34). Journal of Evaluation in Clinical Practice, 15 (4):602–606 | SCNS-SF34 | No multidimensional PREM | Only covers “patient needs” |
| 8 | Fitch, M., & McAndrew, A. | A performance measurement tool for cancer patient information  and satisfaction. Journal of Cancer Education, 2001, 26, 612–618. | N/A | No multidimensional PREM | Only covers patient satisfaction and information |
| 9 | Llewellyn, C. D., Horne, R., McGurk, M., & Weinman, J. | Development and preliminary validation of a new measure to assess satisfaction with information among head and neck cancer patients: the satisfaction with cancer information profile (SCIP). Head and Neck, 2006, 28  (6):540–548 | SCIP | No multidimensional PREM | Only covers patient satisfaction and information |
| 10 | Lo, C., Burman, D., Rodin, G., & Zimmermann, C. | Measuring patient satisfaction inoncology palliative care: psychometric properties of the FAMCARE-patient scale. Quality of Life Research,18 (6), 747–752 | FAMCARE | No generic, surgery- or cancer care-specific PREM | Only palliative care |
| 11 | Teno, J. M., Lima, J. C., & Lyons, K. D. | Cancer patient assessment and reports of excellence: reliability and validity of advanced cancer patient perceptions of the quality of care. Journal of Clinical Oncology, 2009, 27 (10), 1621–1626. | N/A | No generic, surgery- or cancer care-specific PREM | Only palliative care |
| 12 | van Weert, J. C., Jansen, J., de Bruijn, G.-J., Noordman, J., van Dulmen, S., & Bensing, J. M. | QUOTEchemo: a patient-centred instrument to measure quality of communication preceding chemotherapy treatment through the patient’s eyes. European Journal of Cancer, 2009, 45, 2967–2976 | QUOTEchemo | No generic, surgery- or cancer care-specific PREM | Only covery chemotherapy |
| 13 | Wright, E. P., Kiely, M., Johnston, C., Smith, A.B., Cull, A., & Selby, P. J. | Development and evaluation of an instrument to assess social difficulties in routine oncology practice. Qualify of Life Research, 2005, 14 (2):373–386 | N/A | No multidimensional PREM | Only covers „social difficulties“ |
| 14 | Trask, P. C., Tellefsen, C., Espindle, D., Getter, C., & Hsu, M. A. | Psychometric validation  of the cancer therapy satisfaction survey. Value Health, 2008, 11 (4), 669–679. | CTSQ | No multidimensional PREM | Only covers „patient satisfaction“ and „preference for chemotherapy |
| 15 | Harley C, Adams J, Booth L, Selby P, Brown J, Velikova G. | Patient experiences of continuity of cancer care: development of a new medical care questionnaire (MCQ) for oncology outpatients. Value Health. 2009;12(8):1180‐1186 | MCQ | PREM for outpatient sector | Outpatients only |
| 16 | Kleeberg UR, Feyer P, Günther W, Behrens M. | Patient satisfaction in outpatient cancer care: a prospective survey using the PASQOC questionnaire. Support Care Cancer. 2008;16(8):947–54. | PASQOC  (Patient Satisfaction & Quality in Oncological Care) | PREM for outpatient sector | Outpatients only. In German. |
| 17 | Arora, N. K., Reeve, B. B., Hays, R. D., Clauser, S. B., & Oakley-Girvan, I. | Assessment of quality of cancer-related follow-up care from the cancer survivor’s perspective. Journal of Clinical Oncology, 2011, 29 (10), 1280–1289 | APECC  (Assessment of Patients' experience with cancer care) | PREM for outpatient sector | Only outpatient cancer follow-up |
| 18 | Ouwens, M., Hermens, R., Hulscher, M., Vonk-Okhuijsen, S., Tjan-Heijnen, V., Termeer, R., et al. | Development of indicators for patient-centred cancer care. Supportive Care in cancer, 18 (1), 121–130 | N/A | Disease-specific PREM | PREM for non-small cell lung cancer (NSCLC) |
| 19 | Defossez G, Mathoulin‐Pelissier S, Ingrand I, et al. | Satisfaction with care among patients with non‐metastatic breast cancer: development and first steps of validation of the REPERES‐60 questionnaire. BMC Cancer 2007;7:129–139. | REPERES-60 | Disease-specific PREM | PREM for breast cancer |
| 20 | Ouwens, M. M., Marres, H. A., Hermens, R. R., Hulscher, M. M., van den Hoogen, F. J., Grol, R.  P., & Wollersheim, H. C. | Quality of integrated care for patients with head and neck  cancer: development and measurement of clinical indicators. Head and Neck, 2007, 29 (4), 378–386. | N/A | Disease-specific PREM | PREM for head and neck cancers |
| 21 | Damman, O. C., Hendriks, M., & Sixma, H. J. | Towards more patient centred healthcare: a new Consumer Quality Index instrument to assess patients’ experiences with breast care. European Journal of Cancer, 2009, 45 (9), 1569–1577. | N/A | Disease-specific PREM | PREM for breast cancer |
| 22 | Tarrant C, Baker R, Colman AM, et al. | The prostate care questionnaire for patients (PCQ‐P): reliability, validity and acceptability. BMC Health Serv Res 2009;9:199 | PCQ‐P | Disease-specific PREM | PREM for prostate cancer |
| 23 | de Kok M, Sixma HJ, van der Weijden T, et al | A patient‐centred instrument for assessment of quality of breast cancer care: results of a pilot questionnaire. Qual Saf Health Care 2010;19:e40 | QUOTE‐BC | Disease-specific PREM | PREM for breast cancer |
| 24 | Wong WS, Fielding R, Wong C, Hedley A | Confirmatory factor analysis and sample invariance of the Chinese Patient Satisfaction Questionnaire (ChPSQ‐9) among patients with breast and lung cancer. Value Health 2009;12:597–605 | ChPSQ‐9 | Disease-specific PREM | Only for Chinese patients. Only for lung, breast, and hepatocellular carcinoma |
| 25 | Lubeck DP, Litwin MS, Henning JM, Mathias SD, Bloor L, Carroll PR | An instrument to measure patient satisfaction with healthcare in an observational database: results of a validation study using data from CaPSURE. Am J Manag Care 2000;6:70–76. | CaPSURE | Disease-specific PREM | PREM for prostate cancer |
| 26 | Boyes, A., Girgis, A., & Lecathelinais, C. | Brief assessment of adult cancer patients’ perceived needs: development and validation of the 34-item Supportive Care Needs Survey (SCNS-SF34). Journal of Evaluation in Clinical Practice, 2009, 15 (4):602–606 | SCNS-SF34 | No generic, surgery- or cancer care-specific PREM | Covers only „supportive care“ |
| 27 | Fitch, M., & McAndrew, A. | A performance measurement tool for cancer patient information  and satisfaction. Journal of Cancer Education, 2011, 26, 612–618. | N/A | No multidimensional PREM | Covers only patient satisfaction and information |
| 28 | Wright, E. P., Kiely, M., Johnston, C., Smith, A.B., Cull, A., & Selby, P. J. | Development and evaluation of an instrument to assess social difficulties in routine oncology practice. Qualify of Life Research, 14 (2):373–386 | N/A | No multidimensional PREM | Covers only „social difficulties“. |
| 29 | Abbasi-Moghaddam MA, Zarei E, Bagherzadeh R, Dargahi H, Farrokhi P. | Evaluation of service quality from patients' viewpoint. BMC Health Serv Res. 2019;19(1):170 | N/A | No multidimensional PREM | No PREM, but *Health care quality questionnaire*. Iran only.. |
| 30 | Barrios-Ipenza F, Calvo-Mora A, Velicia-Martin F, Criado-Garcia F, Leal-Millan A. | Patient Satisfaction in the Peruvian Health Services: Validation and Application of the HEALTHQUAL Scale. International journal of environmental research and public health. 2020;17(14). | HEALTH-QUAL | PREM for outpatient sector | Only for outpatients |
| 31 | Barth J, Kern A, Luthi S, Witt CM. | Assessment of patients' expectations: development and validation of the Expectation for Treatment Scale (ETS). BMJ Open. 2019;9(6):e026712. | ETS | No multidimensional PREM | Only measures patient expectation. |
| 32 | Baumann E, Kuba K, Gotze H, Mehnert-Theuerkauf A, Esser P. | Initial validation of the German version of the Attentional Function Index in a sample of hematological cancer survivors. European journal of cancer care. 2020;29(4):e13226 | AFI | No multidimensional PREM | Measures „attentional functioning“ only |
| 33 | Martins A, Bennister L, Fern LA, Gerrand C, Onasanya M, Storey L, et al. | Development of a patient-reported experience questionnaire for patients with sarcoma: the Sarcoma Assessment Measure (SAM). Qual Life Res. 2020;29(8):2287-97. | SAM | PROM, no PREM | PROM, no PREM |
| 34 | Hertel-Joergensen M, Abrahamsen C, Jensen C. | Translation, adaptation and psychometric validation of the Good Perioperative Nursing Care Scale (GPNCS) with surgical patients in perioperative care. Int J Orthop Trauma Nurs. 2018;29(101528681):41-8. | GPNCS | No multidimensional PREM | Only covery „perioperative nursing care“ |
| 35 | Alam R, Montanez J, Law S, Lee L, Pecorelli N, Watanabe Y, et al.. | Development of a conceptual framework of recovery after abdominal surgery. Surgical Endoscopy. 2020;34(6):2665-74 | N/A | No multidimensional PREM | Framework for „recovery after surgery“ |
| 36 | Chen S-B, Yang X-Y, Zhang L, Zhang J-Y. | [Validity of the Inpatient Satisfaction Questionnaire]. Sichuan Da Xue Xue Bao Yi Xue Ban. 2018;49(3):425-9 | N/A | No multidimensional PREM | Only measures satisfaction. In Chinese only. |
| 37 | He X, Li L, Bian Y. | Satisfaction survey among primary health care outpatients in the backward region: An empirical study from rural Western China. Patient Preference Adherence. 2018;12((He, Bian) State Key Laboratory of Quality Research in Chinese Medicine, Institute of Chinese Medical Sciences, University of Macau, Taipa, Macao):1989-96. | N/A | PREM for outpatient sector | Only for outpatients Validated only in rural China. |
| 38 | Hussain A, Asif M, Jameel A, Hwang J, Sahito N, Kanwel S. | Promoting OPD patient satisfaction through different healthcare determinants: A study of public sector hospitals. International Journal of Environmental Research and Public Health. 2019;16(19):3719 | N/A | No multidimensional PREM | Only measures satisfaction No validation. |
| 39 | Kilpatrick K, Tchouaket E, Paquette L, Guillemette C, Jabbour M, Desmeules F, et al. | Measuring patient and family perceptions of team processes and outcomes in healthcare teams: questionnaire development and psychometric evaluation. BMC Health Serv Res. 2019;19(1):9 | N/A | No multidimensional PREM | Only measures „team outcomes“ |
| 40 | Lapp V. | The Patient's Voice: Development of an Adolescent Hospital Quality of Care Survey (AHQOCS). J Pediatr Nurs. 2019;49 | AHQOCS | Only for adolescents | Only for adolescents . |
| 41 | Qayyum R, Mubbashir Sheikh M, Uhelski AC, Shaukat F, Shahid M, Panda M. | Comparison of two surveys examining satisfaction of hospitalized patients with physician communication. J Hosp Med. 2018;13(4 Supplement 1). | N/A | No multidimensional PREM | Only measures satisfaction with doctor´s communication |
| 42 | van Melle MA, van Stel HF, Poldervaart JM, de Wit NJ, Zwart DLM. | The transitional risk and incident questionnaire was valid and reliable for measuring transitional patient safety from the patients' perspective. Journal of clinical epidemiology. 2019;105(jce, 8801383):40-9. | N/A | No multidimensional PREM | Only measures „transitional patient safety” |
| 43 | Wong EL-Y, Cheung AW-L, Xu RH, Yam CH-K, Lui S-F, Yeoh E-K. | Development and validation of a generic patient experience instrument for measuring specialist outpatient service in Hong Kong. Int J Qual Health Care. 2019;31(10):G158-G64 | N/A | PREM for outpatient sector | Only for outpatients |
| 44 | Wu Y, Mu J, Zhang S. | Evaluating Patient Satisfaction in Township Hospitals in the Cold Regions of China. HERD. 2020((Wu, Mu, Zhang) Key Laboratory of Cold Region Urban and Rural Human Settlement Environment Science and Technology, School of Architecture, 47822Harbin Institute of Technology, China):1937586720958016. | N/A | No multidimensional PREM | Only measures satisfaction |
| 45 | Zhang J, Zhou F, Ge X, Ran X, Li Y, Chen S, et al. | Reliability and validity of an indicator system used to evaluate outpatient and inpatient satisfaction in Chinese hospitals. Patient Preference Adherence. 2018;12((Zhang) Department of Neurology, Peking University People's Hospital, Beijing, China):2527-36 | N/A | No multidimensional PREM | Only measures satisfaction. In Chinese. |
| 46 | Ziabakhsh S, Albert A, Houlihan E. | Development and Validation of a Brief Hospital-Based Ambulatory Patient Experience Survey Tool. Healtc Policy. 2019;15(2):E100-E14 | N/A | PREM for outpatient sector | Only for outpatients |
| 47 | Albornoz-Cabello M, Perez-Marmol JM, Cardero-Duran MA, Barrios-Quinta CJ, Espejo-Antunez L. | Construction, factor structure, and internal consistency reliability of the hospital physical therapy perceived satisfaction questionnaire (H-ptps). International Journal of Environmental Research and Public Health. 2020;17(16):1-14 | H-ptps | No multidimensional PREM | Only measures satisfaction. Only for rehabilitation. |
| 48 | Baldie DJ, Guthrie B, Entwistle V, Kroll T. | Exploring the impact and use of patients' feedback about their care experiences in general practice settings-A realist synthesis. Family Practice. 2018;35(1):13-21 | N/A | PREM for outpatient sector | PREM for general practitioners |
| 49 | Beckers E, Webers C, Boonen A, ten Klooster PM, Vonkeman HE, van Tubergen A. | Validation and implementation of a patient-reported experience measure for patients with rheumatoid arthritis and spondyloarthritis in the Netherlands. Clinical Rheumatology. 2020;39(10):2889-97 | N/A | Disease-specific PREM | PREM for rheumatoide Arthritis |
| 50 | Berzina-Novikova N, Taube M. | Patient satisfaction questionnaire adaptation process-experience from Latvia. Eur Psychiatry. 2018;48(Supplement 1):S444 | PIPES | Disease-specific PREM | PREM for psychiatric diseases |
| 51 | Borg S, Eeg-Olofsson K, Palaszewski B, Svedbo Engstrom M, Gerdtham UG, Gudbjornsdottir S. | Patient-reported outcome and experience measures for diabetes: Development of scale models, differences between patient groups and relationships with cardiovascular and diabetes complication risk factors, in a combined registry and survey study in Sweden. BMJ Open. 2019;9(1):e025033. | N/A | Disease-specific PREM | PREM for Diabetes mellitus |
| 52 | Grundy A, Keetharuth AD, Barber R, Carlton J, Connell J, Taylor Buck E, et al. | Public involvement in health outcomes research: Lessons learnt from the development of the recovering quality of life (ReQoL) measures. Health and Quality of Life Outcomes. 2019;17(1):60. | ReQoL | Disease-specific PREM | PREM for psychiatric diseases |
| 53 | Husebo AML, Morken IM, Eriksen KS, Nordfonn OK. | The patient experience with treatment and self-management (PETS) questionnaire: translation and cultural adaption of the Norwegian version. BMC Med Res Methodol. 2018;18(1):147 | PETS | Disease-specific PREM | PREM for chronic diseases |
| 54 | Kimman ML, Wijsenbeek MS, van Kuijk SMJ, Wijnsma KL, van de Kar NCAJ, Storm M, et al. | Validity of the Patient Experiences and Satisfaction with Medications (PESaM) Questionnaire. Patient. 2019;12(1):149-62 | PESaM | No multidimensional PREM | Patient Experiences and Satisfaction with Medications |
| 55 | Kremers MNT, Mols EEM, Simons YAE, van Kuijk SMJ, Holleman F, Nanayakkara PWB, et al. | Quality of acute internal medicine: A patient-centered approach. Validation and usage of the Patient Reported Measure-acute care in the Netherlands. PLoS ONE. 2020;15(12 December):e0242603 | N/A | Disease-specific PREM | PREM for „acute care“ |
| 56 | Luther L, Fukui S, Garabrant JM, Rollins AL, Morse G, Henry N, et al. | Measuring Quality of Care in Community Mental Health: Validation of Concordant Clinician and Client Quality-of-Care Scales. J Behav Health Serv Res. 2019;46(1):64-79 | N/A | Disease-specific PREM | PREM for „community mental health” |
| 57 | Pap R, Lockwood C, Stephenson M, Simpson P. | Indicators to measure prehospital care quality: A scoping review. JBI Database Syst Rev Implement Rep. 2018;16(11):2192-223. | N/A | No multidimensional PREM | Scoping Review for a prehospital care PREM |
| 58 | Eskildsen NB, Ross L, Bulsara C, Dietz SM, Thomsen TG, Groenvold M, et al. | Development and content validation of a questionnaire measuring patient empowerment in cancer follow-up. Qual Life Res. 2020;29(8):2253-74 | N/A | No multidimensional PREM | *patient-reported empowerment* questionnaire |
