## Supplementary material for "Measuring patient centeredness with German language Patient-Reported Experience Measures (PREM) – a systematic review and qualitative analysis according to COSMIN": S1_Figure

### Slide 1
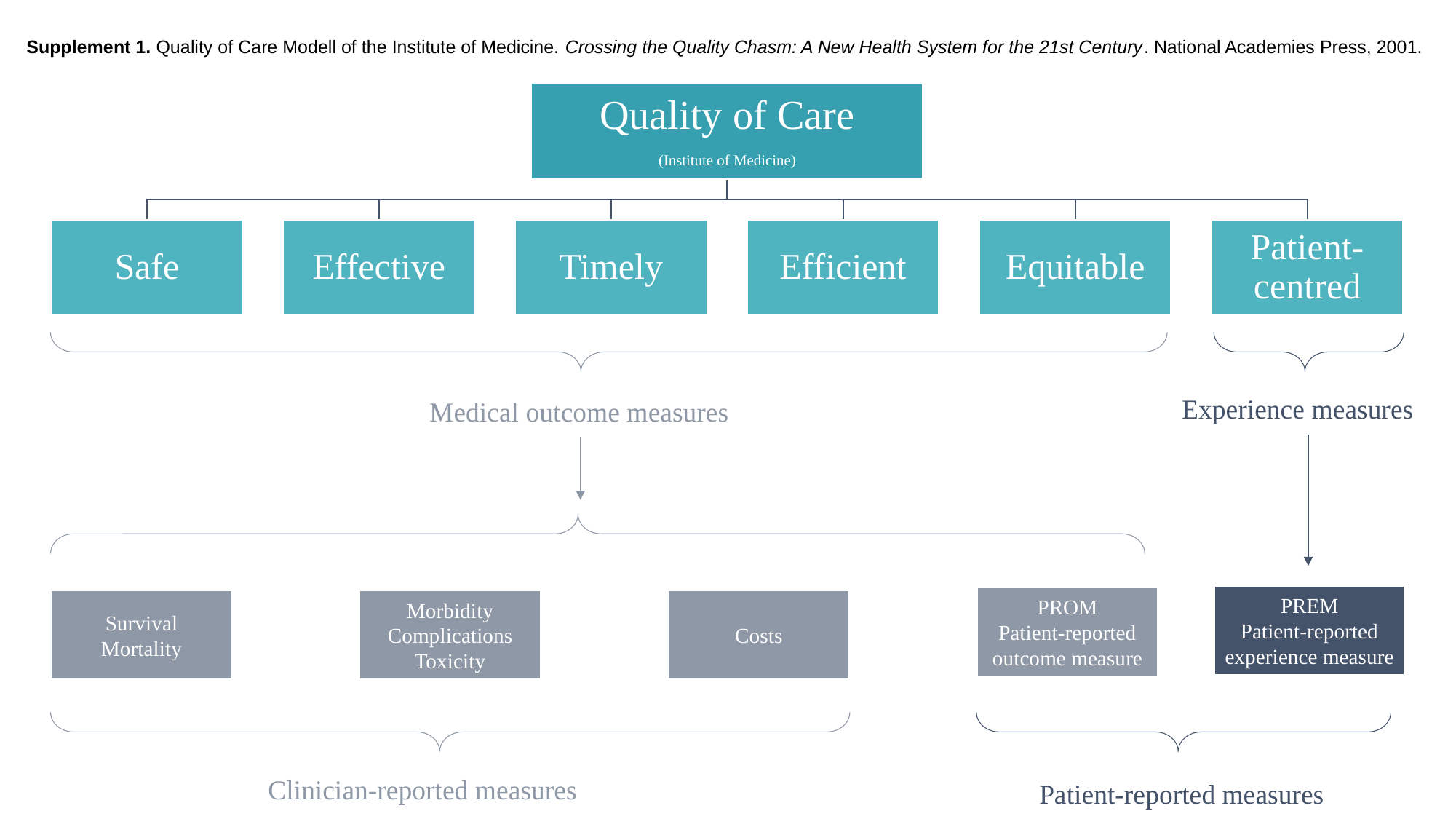

Supplement 1. Quality of Care Modell of the Institute of Medicine. Crossing the Quality Chasm: A New Health System for the 21st Century. National Academies Press, 2001.
Experience measures
Medical outcome measures
PREM
Patient-reported experience measure
PROM
Patient-reported outcome measure
Survival
Mortality
Morbidity
Complications
Toxicity
Costs
Clinician-reported measures
Patient-reported measures
